# Leveraging Deep Neural Network and Language Models for Predicting Long-Term Hospitalization Risk in Schizophrenia

**DOI:** 10.1101/2024.11.27.24317896

**Authors:** Yihang Bao, Wanying Wang, Zhe Liu, Weidi Wang, Xue Zhao, Shunying Yu, Guan Ning Lin

## Abstract

Early warning of long-term hospitalization in schizophrenia (SCZ) patients at the time of admission is crucial for effective resource allocation and individual treatment planning. In this study, we developed a deep learning model that integrates demographic, behavioral, and blood test data from admission to forecast extended hospital stays using a retrospective cohort. By utilizing language models (LMs), our developed algorithm efficiently extracts 95% of the unstructured electronic health record data needed for this work, while ensuring data privacy and low error rate. This paradigm has also been demonstrated to have significant advantages in reducing potential discrimination and erroneous dependencies. By utilizing multimodal features, our deep learning model achieved a classification accuracy of 0.81 and an AUC of 0.9. Key risk factors identified included advanced age, longer disease duration, and blood markers such as elevated neutrophil-to-lymphocyte ratio, lower lymphocyte percentage, and reduced albumin levels, validated through comprehensive interpretability analyses and ablation studies. The inclusion of multimodal data significantly improved prediction performance, with demographic variables alone achieving an accuracy of 0.73, which increased to 0.81 with the addition of behavioral and blood test data. Our approach outperformed traditional machine learning methods, which were less effective in predicting long-term stays. This study demonstrates the potential of integrating diverse data types for enhanced predictive accuracy in mental health care, providing a robust framework for early intervention and personalized treatment in schizophrenia management.

## Main

Schizophrenia (SCZ) is a chronic and severe mental disorder that profoundly affects an individual’s thoughts, feelings, and behaviors. Patients with SCZ often experience symptoms such as delusions, hallucinations, disorganized thinking, and impaired functioning, leading to significant distress and disability [1, 2]. These symptoms pose substantial risks not only to the patients but also to society at large. Due to the increased risk of suicide and other health problems, individuals with SCZ have a higher mortality rate, resulting in a reduced lifespan of approximately 15-20 years [3]. Given these risks, inpatient treatment is crucial for effectively managing and treating individuals with SCZ [4]. However, the rising prevalence of SCZ [5], coupled with a large patient population, exacerbates the challenges related to the allocation of healthcare resources [6]. Healthcare institutions must allocate resources based on the individual conditions of patients upon admission to enhance treatment efficiency and ensure early intervention for those at risk of treatment resistance. A critical aspect of this process is the reliable early identification of patients at risk of prolonged hospitalization and the understanding the factors that significantly impact the length of stay [7].

Long-stay patients, defined as those requiring hospitalization for an extended period due to the severity and complexity of their condition, present a significant challenge for healthcare systems. Identifying these patients at the time of admission is crucial. Studies have shown that early identification of long-stay patients can enhance treatment efficiency and reduce the financial burden of patients [8–11]. For instance, a study on post-admission medical interventions for COVID-19 patients demonstrated that targeted interventions based on specific patient risk factors could significantly shorten hospital stays [12]. Similarly, a retrospective study in the Philippines indicated that inappropriate allocation of medical resources at the time of admission led to prolonged hospitalizations and increased financial burden for some patients [8]. Thus, early identification of long-stay patients can facilitate better allocation of resources, improve efficiency, and reduce costs from the perspective of healthcare institutions [13].

Hospitalization durations for SCZ are notably long. For example, in South Korea, the average hospitalization duration is 78 ± 76 days [14], in China, it is 73.3 ± 42.2 days [15], in Israel it is 90–120 days [16], and in Canada it is 96.6 days [17]. This highlights the importance of early identification of long-stay patients. To address this need, studies have explored the differences between long-stay and short-stay SCZ patients at the time of admission. Demographic variables, such as age, gender, marital status, employment, and race have been found to influence the length of hospital stay [13, 18–20]. Additionally, behavioral variables, such as the presence of aggressive behavior or suicidal ideation, have been identified as factors affecting hospitalization duration [21–23]. Both types of variables can be obtained at the time of patient admission. Research has also explored the association between clinical variables, such as discharge medication dosage, and the length of hospital stay [19, 24]. However, these clinical variables are not available at the time of admission. Building on these insights, researchers have developed computational models to predict long-stay patients using multi-perspective admission information. For instance, Kirchebner et al., combined patient characteristics, criminal history, and medical history to construct a machine learning based forensic psychiatry hospitalization duration prediction model [25]. Similarly, Cheng et al. developed a regression model for predicting hospitalization duration using demographic and biochemical variables, finding an association between marital status, thyroid-stimulating hormone levels, and prolonged hospital stays [26]. While these studies have made valuable contributions, they also faced several challenges. The study by Kirchebner et al. employed manual methods to extract unstructured data, which are not applicable to large-scale datasets. The work by Cheng et al. was limited by the number of features and algorithm efficiency, resulting in a model with an AUC of only 0.577, thereby reducing its applicability. In light of this, there is a need for an advanced algorithmic framework that combines automatic unstructured data extraction to achieve more accurate long-term hospitalization risk prediction for SCZ patients.

The rapid development of deep neural networks and LMs has enabled more efficient and feasible feature extraction and learning. In this study, we leveraged these advanced techniques to predict long-stay risks in SCZ patients based on various types of admission variables. Specifically, we extracted unstructured behavioral features from a large set of retrospective electronic medical records of SCZ patients using a semi-automated process based on LMs. This extraction process can be quickly completed in a few-shot setting, outperforming methods based on regular expressions or BERT [27, 28], an advanced LMs that uses bidirectional pre-training to thoroughly understand the context of words in sentences. By integrating behavioral features, demographic characteristics, and admission blood test indicators as inputs, our deep neural network successfully learned the differences between short-stay and long-stay patients, and achieved a classification accuracy of 0.81 and an AUC of 0.9 on an independent test set. Furthermore, through interpretability analysis and case studies, we identified key differentiating features such as admission age, disease duration, and marital status, which corroborate previous research findings [16, 18–20]. We also demonstrated the effective contribution of admission blood test indicators to the prediction of long hospital stays for SCZ patients.

## Results

### Patient characteristics and basic analysis

Our retrospective dataset, sourced from the Shanghai Mental Health Center (SMHC), comprises electronic medical records of SCZ inpatients discharged between 2018 and 2022. Details on the data processing and screening methods are shown in the Methods section and Extended Data Figure 1. Based on previous research [15], we defined hospitalization durations exceeding 120 days as long-stay and durations shorter than 30 days as short-stay. Patients with a hospital stay longer than one year were excluded, as their length of stay is typically influenced by non-medical factors. This yielded a dataset of 1,546 short-stay records and 812 long-stay records, along with their corresponding 30-dimensional admission features (see Table 1). These features were categorized into behavioral variables, demographic variables, and admission blood test indicators. Additionally, based on prior researches linking inflammatory factors with SCZ [29, 30], we included calculated inflammatory markers in the blood test indicators. These markers include the Neutrophil-to-Lymphocyte Ratio (NLR), Monocyte-to-Lymphocyte Ratio (MLR), Platelet-to-Lymphocyte Ratio (PLR), and Systemic Immune-Inflammation Index (SII).

**Table 1.** Descriptive statistics on patients.

| LOS | Short | Long | P-value |
| --- | --- | --- | --- |
| N | 1546 | 812 |  |
| Age of admission (years) | $35.9 \pm 18.0$ | $48.7 \pm 17.3$ | $< 1e^{-6}$ |
| Disease duration (days) | $3802 \pm 5016$ | $7932.1 \pm 5651.6$ | $< 1e^{-6}$ |
| Sex |  |  | 0.026 |
| Female | 866(56.0%) | 415(51.1%) |  |
| Male | 680(44.0%) | 397(48.9%) |  |
| Marital status | | | $< 1e^{-6}$ |
| Married | 468(30.3%) | 179(22.0%) |  |
| Divorced/Widowed | 116(7.5%) | 117(14.4%) |  |
| Unmarried | 962(62.2%) | 516(63.5%) |  |
| Hemoglobin | $135.3 \pm 17.6$ | $132.3 \pm 17.3$ | $4.6e^{-5}$ |
| Neutrophil | $4.5 \pm 1.9$ | $4.4 \pm 1.8$ | 0.99 |
| Neutrophil% | $61 \pm 11.5$ | $63.1 \pm 10.4$ | $1.2e^{-5}$ |
| Eosinophil | $0.125 \pm 0.137$ | $0.12 \pm 0.14$ | 0.071 |
| Eosinophil% | $1.87 \pm 2.06$ | $1.8 \pm 2.1$ | 0.45 |
| Basophil | $0.026 \pm 0.017$ | $0.024 \pm 0.018$ | 0.022 |
| Basophil% | $0.38 \pm 0.25$ | $0.37 \pm 0.25$ | 0.25 |
| Lymphocyte | $2.0 \pm 0.7$ | $1.8 \pm 0.7$ | $< 1e^{-6}$ |
| Lymphocyte% | $29.4 \pm 10.4$ | $27.4 \pm 9.3$ | $1.8e^{-5}$ |
| Monocyte | $0.5 \pm 0.2$ | $0.5 \pm 0.2$ | $6.1e^{-4}$ |
| Monocyte% | $7.4 \pm 1.9$ | $7. \pm 2.1$ | 0.34 |
| Platelet | $265.7 \pm 73.6$ | $253.6 \pm 74.5$ | $1.7e^{-4}$ |
| C-reactive protein | $4.8 \pm 12.1$ | $5.3 \pm 11.3$ | $3.1e^{-5}$ |
| Albumin | $43.0 \pm 4.2$ | $41.2 \pm 4.0$ | $< 1e^{-6}$ |
| Globulin | $28.5 \pm 4.6$ | $28.5 \pm 4.7$ | 0.88 |
| Total bilirubin | $14.9 \pm 7.7$ | $13.4 \pm 6.6$ | $3.1e^{-6}$ |
| Alanine aminotransferase | $24.4 \pm 22.5$ | $24.6 \pm 21.2$ | 0.048 |
| Alkaline phosphatase | $84.8 \pm 53.3$ | $77.4 \pm 26.6$ | 0.67 |
| γ-glutamyl transferase | $24.2 \pm 28.3$ | $26.3 \pm 32.7$ | 0.17 |
| Natrium | $140.8 \pm 2.8$ | $141.2 \pm 3.0$ | $5.4e^{-4}$ |
| Kalium | $3.9 \pm 0.6$ | $3.8 \pm 0.4$ | $3.5e^{-3}$ |
| **Chlorine** | $104.9 \pm 3.1$ | $105.5 \pm 3.2$ | $1.1e^{-6}$ |
| **NLR** | $2.7 \pm 2.0$ | $2.9 \pm 2.2$ | $1.3e^{-5}$ |
| **MLR** | $0.3 \pm 0.2$ | $0.3 \pm 0.2$ | 0.019 |
| **PLR** | $148.4 \pm 66.3$ | $160.1 \pm 98.0$ | $4.9e^{-4}$ |
| **SII** | $697.3 \pm 526.0$ | $737.1 \pm 623.9$ | 0.041 |

For these features, we performed statistical analysis, significance testing, and correlation analysis, as detailed in Table 1 and Extended Data Figures 2-4. Continuous variables were analyzed using the Mann-Whitney U test, while categorical variables were assessed using the chi-square test for homogeneity. The results revealed significant univariate associations between admission age (p-value < 1*e*^-6^), disease duration (p-value < 1*e*^-6^), and long-stay hospitalization in the demographic data, corroborating previous research findings [31]. Other significant variables, such as total bilirubin (p-value = 3*e*^-6^) and lymphocytes (p-value < 1*e*^-6^), also have research support for their associations with SCZ [32, 33]. Through correlation analysis, we observed that apart from variables with direct computational relationships, the remaining variables exhibited low correlations with each other (average Absolute Pearson Correlation Coefficient < 0.16). This indicates the diversity of perspectives offered by these features.

### Optimizing Language Models for Confidential Unstructured Data Extraction

Recent advancements in LMs such as ChatGPT [34] have demonstrated performance on par with state-of-the-art supervised learning methods across various natural language processing tasks, such as question answering [35], entity recognition [36], and relation extraction [37]. These LMs offered significant advantage over traditional supervised methods, including the ability to perform few-shot learning or even zero-shot learning capabilities and enhanced interpretability [38]. In our study, we aimed to identify behavioral variables, including aggression, self-destruction, and elopement, from unstructured text. The detailed definitions of the behavioral variables can be found in Extended Data Note. Given the unstructured format nature of EHR data, employing LM-based methods presents a viable solution for addressing the unstructured-text classification challenges.

However, due to stringent confidentiality policies, we could not directly apply state-of-the-art models like ChatGPT. Conversely, locally deployed models often suffered from performance constraints. To circumvent this, we proposed a prompt optimization method using LMs for unstructured information extraction, ensuring data remained fully localized. Detailed implementation steps are outlined in the Method section. In brief, we assessed the performance of local LMs on a small validation set and iteratively optimized the prompts using advanced LMs or manual adjustments based on evaluation results. The workflow and examples of optimized prompts are illustrated in Figures 1a and 1b. evaluation results during the optimization process, depicted in Figures 1c to 1e and Supplementary Table 1, showed that our iterative strategy enables the prompts to achieve within approximately five rounds on a validation set of 100 samples. Notably, Figure 1d exhibited a performance fluctuation, attributed to GPT-4 occasionally following incorrect optimization paths due to the lack of specific data knowledge. Once this decline was identified, the optimization direction was promptly corrected.

**Figure 1.**
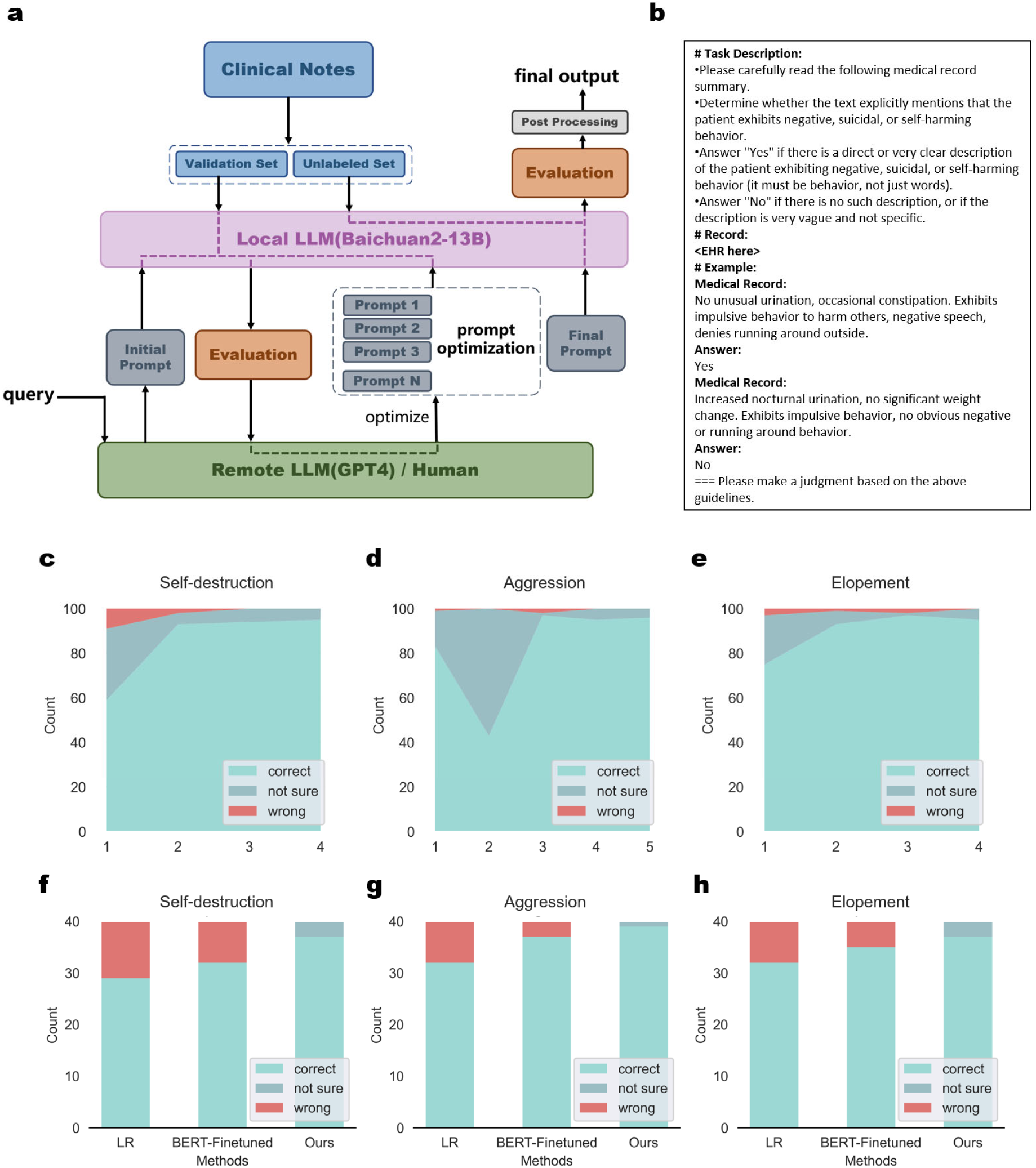
**(a)** Workflow of our few-shot unstructured information extraction method based on local LM. **(b)** Typical structure of the final optimized prompt in this study. **(c-e)** Accuracy on the validation set during prompt optimization. **(f-h)** The comparation of our methods with linear regression and BERT on independent test set. Results show that our methods can achieve the lowest error rate.

To validate the superiority of our method over supervised learning approaches, we manually labeled 40 records as a test set. The comparative results, shown in Figures 1f to 1h and Supplementary Table 2, highlighted our method’s performance against Linear Regression and fine-tuned BERT [27] on tasks including aggression recognition, self-destruction recognition, and elopement recognition. Specifically, both supervised learning models were pre-trained on the validation set before being tested on the labeled test set. Our method not only achieved the highest accuracy (average 94.2%) but also identified records where the LM could not provide a judgment. These few uncertain records can be manually annotated, ensuring high accuracy while significantly reducing the manual workload.

In summary, our information extraction method based on LMs utilizes iteratively optimized prompts to extract data while maintaining data confidentiality. Our approach, validated through multiple perspectives, significantly reduced manual workload and achieved near-perfect accuracy in extracting behavioral variables, demonstrating the efficacy of LMs in unstructured information extraction.

### Predicting Long-Term Hospitalization Risk in SCZ Patients Using Integrated Demographic, Behavioral, and Blood Test Indicators

We hypothesized that integrating demographic variables, behavioral variables, and blood test indicators would enhance the prediction of long-term hospitalization risk in SCZ patients. The workflow of our model is depicted in Figure 2a. Structured demographic variables and blood test indicators underwent preprocessing operations, including normalization and variable transformation. For unstructured data, we utilized an LM-based information extraction strategy. These features were consolidated into a 33-dimensional vector and fed into an MLP network architecture with attention mechanisms for feature extraction and binary classification of long and short hospitalizations. Comprehensive details on data preprocessing and the model process are available in the methods section.

**Figure 2.**
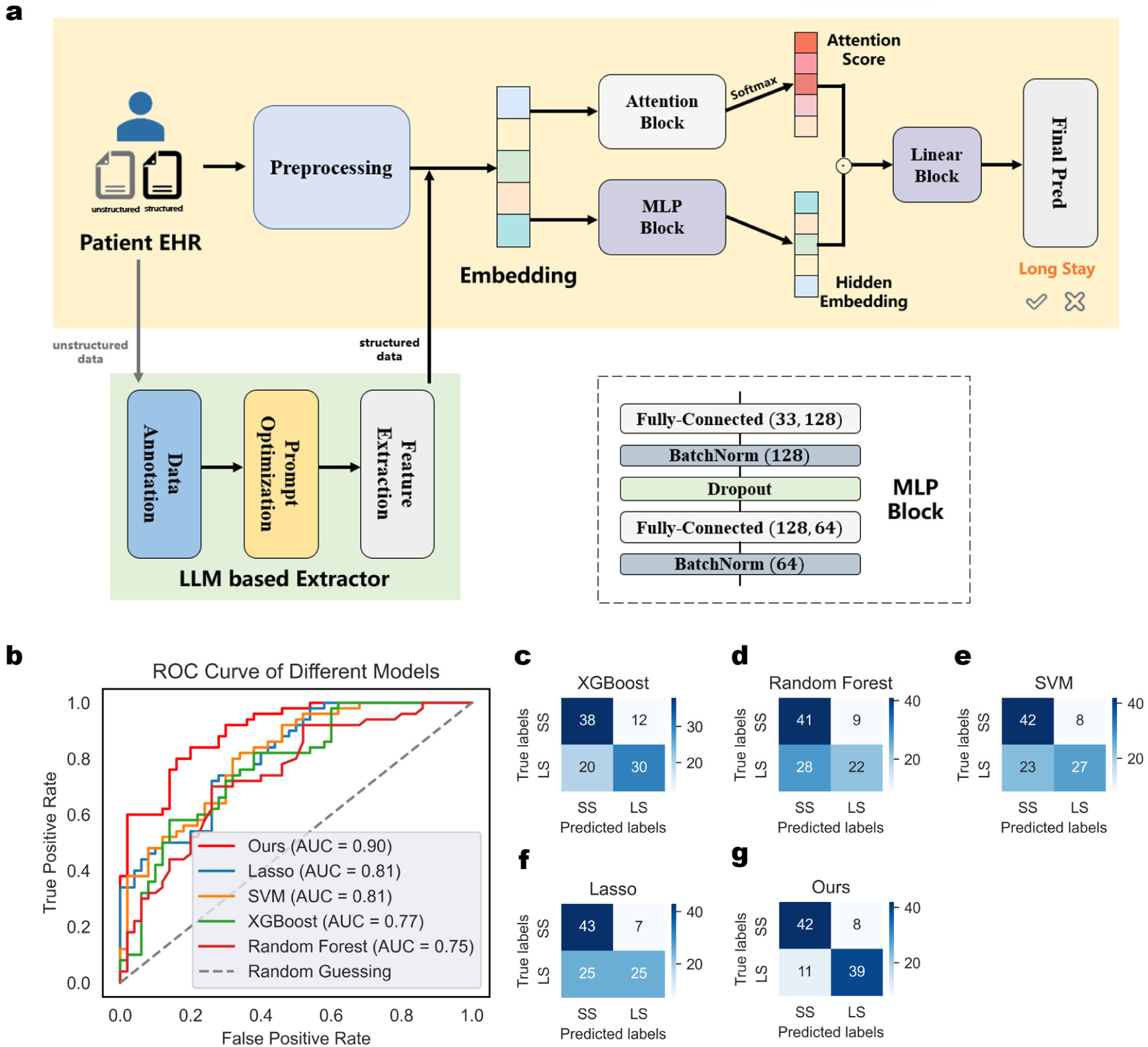
**(a)** Architecture of our long-term hospitalization risk prediction model **(b)** ROC curve of our model and machine learning methods on test set **(c-g)** Confusion matrix of our model and machine learning methods on test set

During training, we applied leave-one-out cross-validation [39] to optimize our hyperparameters. Post-optimization, we evaluated our model’s performance on an independent test set and benchmarked it against other machine learning methods. The independent test set, partitioned prior to training, comprised the most recent 50 long-term and short-term hospitalization patients from the dataset. This temporal segmentation strategy, grounded in the latest data, has been validated as both effective and reasonable [40]. Figure 2b illustrated the ROC curves of our model versus other machine learning methods on the independent test set, while Figures 2c-g display the corresponding confusion matrices. Precision, recall, and other metrics derived from the confusion matrices are detailed in Supplementary Table 3. Our model achieved an AUC of 0.9 and an accuracy of 0.81, demonstrating its efficacy in predicting long-term hospitalization risk for SCZ patients based on multifaceted admission variables. By analyzing the confusion matrices of each model, we observed that conventional machine learning models well in predicting accuracy for short-stay patients, their predictions for long-stay patients were close to random guessing. Conversely, our model effectively identified the characteristics of long-stay patients, demonstrating a marked improvement over traditional methods.

### Comprehensive interpretability analysis revealed associations between admission variables and prolonged hospitalization for SCZ

We hypothesized that a comprehensive interpretability analysis of our black-box model would not only validate its effectiveness but also elucidate the admission risk factors associated with prolonged hospitalization for SCZ. Firstly, we conducted representation visualization using Principal Component Analysis to reduce the dimension of both input vectors and final hidden vectors to two dimensions, as shown in Figures 3a and 3b. The projections of the original input vectors in the two-dimensional space did not form discernible clusters. In contrast, the final hidden vectors computed by the model exhibited distinct classification boundaries, indicating that our model effectively identified a hyperplane that separated the dataset.

**Figure 3.**
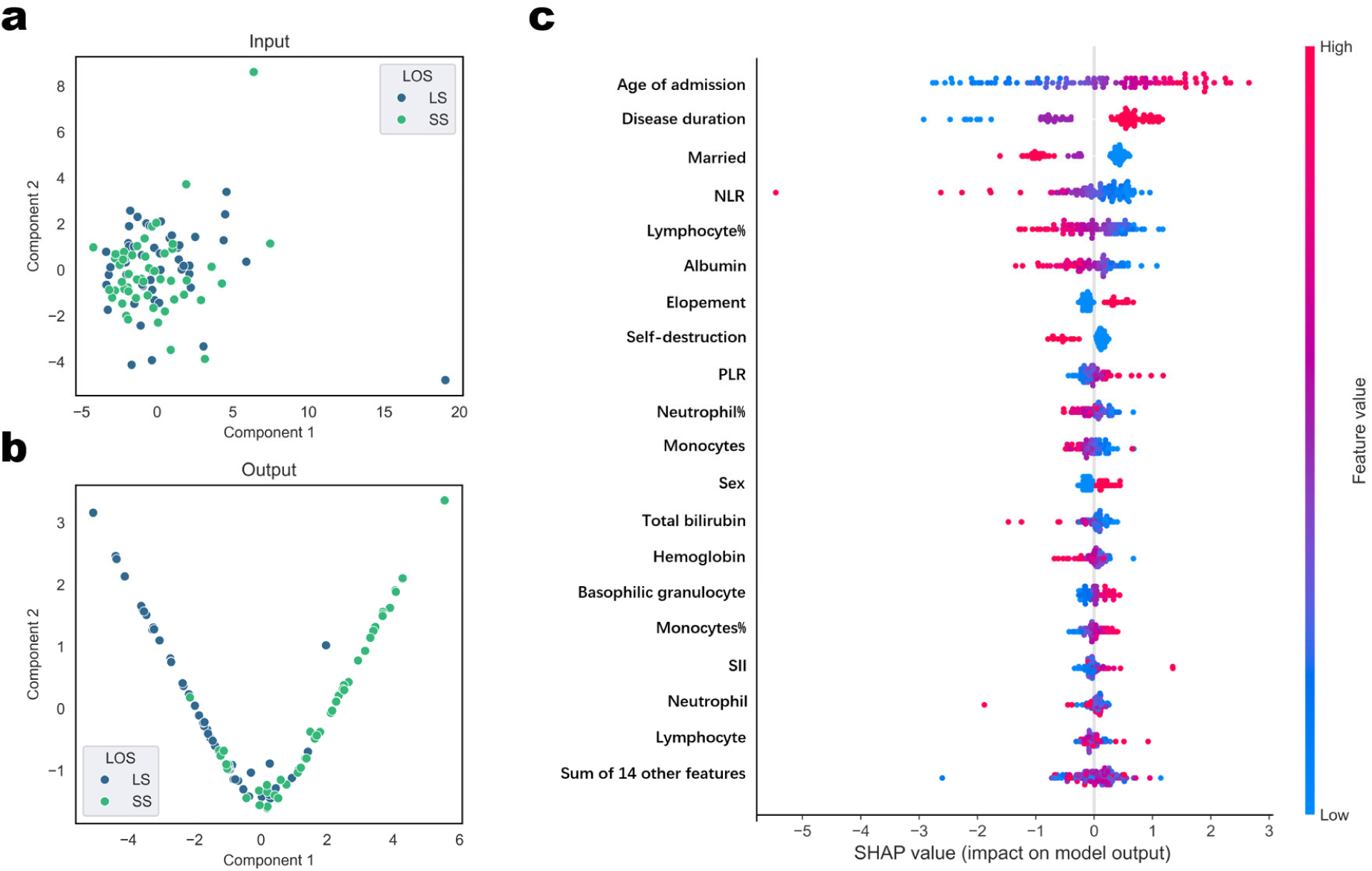
**(a-b)** Interpretability analysis performed on a single patient using SHAP. **(c)** Model interpretability analysis on test set using SHAP. The color bar corresponds to the raw values of the variables for each instance. In this beeswarm plot, points are distributed horizontally along the x-axis according to their SHAP value.

To further dissected the significant features in our classification process, we employed SHapley Additive exPlanations (SHAP) for model interpretability analysis [41]. SHAP provides a unified approach to interpreting machine learning models by calculating the average marginal contribution of each feature value over all possible coalitions. Figures 3b and Extended Data Figures 5-6 illustrate the SHAP analysis results on our test set. The colors in the figures represent the values of the corresponding features, and the x-axis denotes the SHAP values of the features. Key observations included: 1) Demographic Variables: Age of Admission (positively correlated), Disease Duration (positively correlated), and Marital Status (negatively correlated) emerged as three critical risk factors for prolonged hospitalization, aligning with previous studies [20, 26, 42]; 2) Blood Test Indicators: Blood test indicators such as inflammatory markers, Lymphocyte%, and Albumin were significant. Previous studies had linked these indicators with the severity of SCZ symptoms [43] and the effectiveness of medication [44], and this work is the first to associate them with prolonged hospitalization; 3) Behavioral Variables: Behavioral variables like elopement and self-destructive behaviors also correlated with extended hospital stays.

### Ablation study and case analysis revealed the effectiveness of multi-perspective variables in aiding long-term hospitalization risk prediction

To illustrate the effectiveness of using multi-perspective variables, we conducted ablation experiments by incrementally adding blood test indicators and behavioral variables to the demographic variables. The detailed procedure is described in the Methods section. Figures 4b-e and Supplementary Table 4 present the results of these experiments. Using demographic variables alone achieved an AUC close to 0.8, demonstrating their strong predictive foundation. Adding blood test indicators and behavioral variables significantly improved model performance to an AUC of 0.85 and 0.83, with the combined inputs of all three types of variables further enhancing the performance to match the original model with an AUC of 0.9. This indicated that while demographic variables provide a solid predictive base, the inclusion of blood test indicators and behavioral variables adds additional, valuable information, improving the prediction of long-term hospitalization risk by an AUC of 6.25% and 3.75%. Furthermore, the examination of confusion matrices revealed that introducing blood test indicators significantly reduced the false positive rate by 31% (from 0.32 to 0.22). This is likely due to the direct health status information these indicators provided, which reduced the likelihood of misclassification. Additionally, we explored how our information extraction paradigm contributes to this research by replacing the behavioral variables extracted by our LM with those extracted using BERT. We observed a decline in model performance (from 0.901 to 0.882). The modest decrease may be attributed to the fact that behavioral variables do not dominate the predictive power in this context; however, the advantages of using our extraction paradigm will become more pronounced when applied to other dominant features in information extraction tasks.

**Figure 4.**
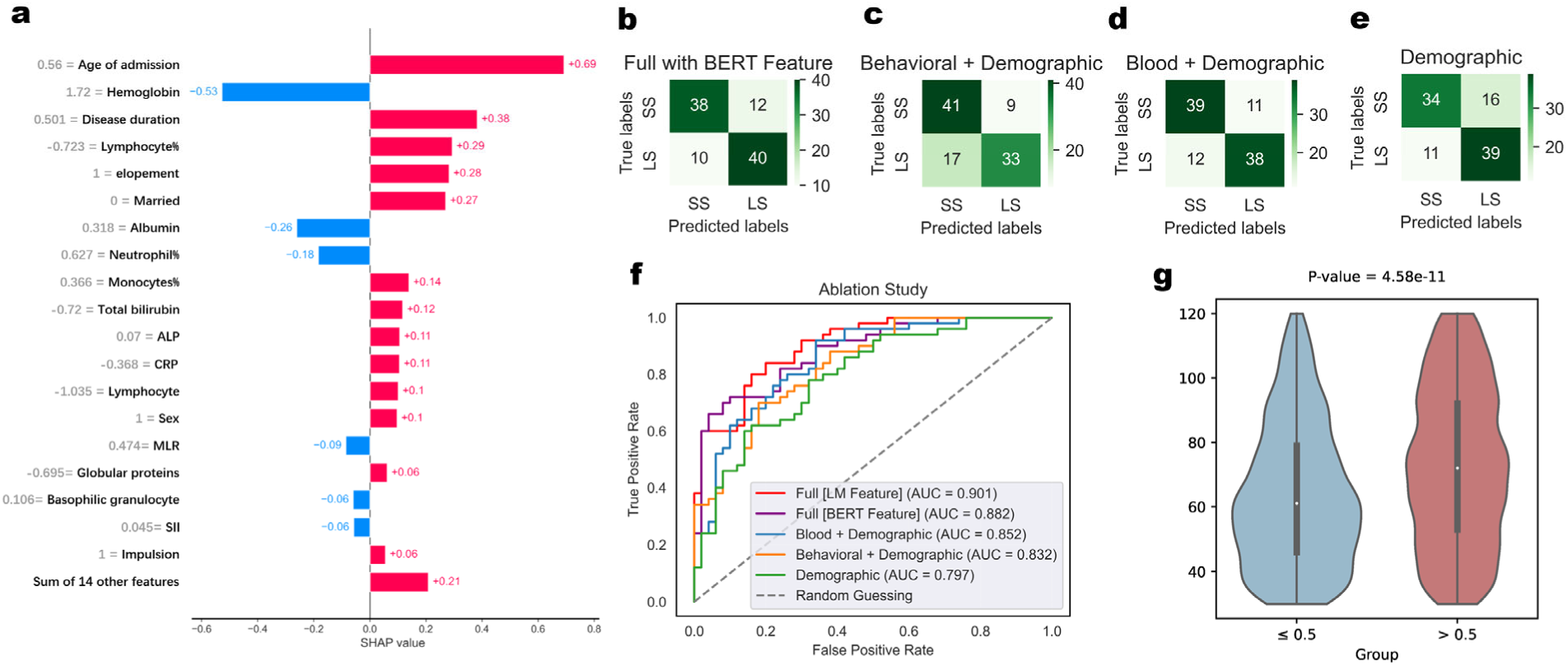
**(a)** Interpretability analysis performed on a single patient using SHAP **(b)** Confusion matrix of the models with BERT extracted behavioral features **(c-e)** Confusion matrix of the models with different data variables were ablated **(f)** ROC curve of the models with different data variables was ablated **(g)** Patients with intermediate length of stay were grouped according to the model prediction. The significant differences shown in the figure illustrate the effectiveness of our model.

To further illustrate the importance of multi-perspective variables in predicting long-term hospitalization from an individual perspective, we conducted a case study analyzing SHAP values for a specific patient—a male with a hospitalization duration of 161 days. Age and disease duration were found to have the most significant impact on predicting hospitalization duration, consistent with previous studies [45, 46] that correlated older age with longer hospital stays. SCZ, as a severe chronic illness, often involves long disease durations, correlating with recovery difficulty and extended hospitalization periods. Reviewing the EHR revealed poor medication adherence, strongly associated with condition severity and longer hospital stays. In addition, aggressive and elopement behaviors noted in the records indicated poor adherence. Blood test indicators related to blood cells, liver function, and electrolytes supported the prediction of prolonged hospitalization, as detailed in the Extended Data Note. This case study demonstrated that incorporating multi-perspective variables not only enhances the accuracy and comprehensiveness of the prediction but also allowed for a multi-faceted analysis of patient conditions upon admission. This approach facilitated more efficient allocation of medical resources and the development of tailored treatment plans.

### Our information extraction paradigm exhibited superiority in enhancing accuracy, reducing information bias, and minimizing erroneous information dependency

To demonstrate the superiority of our LM extraction paradigm, we conducted a multi-faceted study. First, to validate the effectiveness of our paradigm on external cohorts, we performed tests on an external synthetic dataset. The results, shown in Figure 5a, indicate that our method maintains optimal accuracy, achieving a 95% correct rate. From another perspective, we assessed the performance of our approach in the presence of biases. We introduced two bias scenarios: racial bias and gender bias. Detailed testing methodologies can be found in the Methods section, with results illustrated in Figures 5b-d and Extended Data Figures 8-9. Notably, supervised learning methods exhibited a significant performance decline in the presence of bias (Wilcoxon test, P=0.0045), whereas our method remained unaffected. This indicates that our approach effectively mitigates potential unfairness related to racial and gender discrimination. Additionally, we considered another scenario involving erroneous information dependencies, where the model might focus on variables unrelated to our actual conclusions. For instance, when extracting whether a patient has a history of self-harm, it is inappropriate to infer this based on age or gender, even if strong correlations exist. We constructed such a scenario by inserting correlated information based on labels to influence the information extraction task. The results, presented in Figure 5e and Extended Data Figure 10, show that supervised models were adversely affected by the misleading correlated information, incorrectly using age as a characteristic to determine self-harming behavior, while our method maintained performance comparable to the original task.

**Figure 5.**
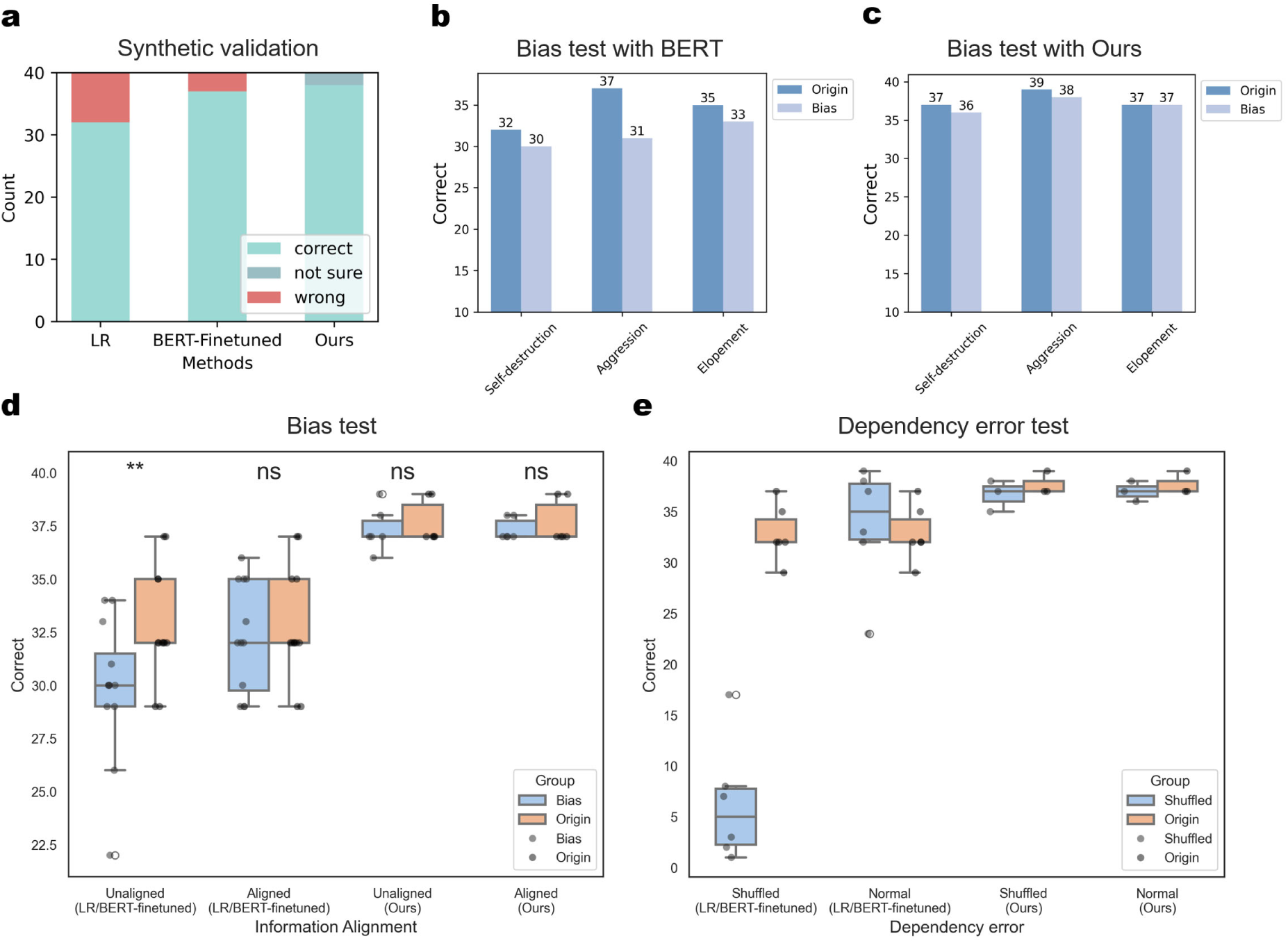
**(a)** The comparation of our methods with linear regression and BERT on synthetic test set. Results show that our methods can achieve the lowest error rate. (b) BERT model performance on original and racial bias datasets. (c) Our model performance on original and racial bias datasets. (d) A study on the performance degradation of the model on racial bias and gender bias datasets shows significant performance decline of the supervised model, as indicated by the Wilcoxon test (p=0.0045). (e) A study on the performance degradation of the model on dependency error datasets shows significant performance decline of the supervised model.

### Correlation analysis in the population with intermediate length of stay

In addition to predicting the hospitalization duration for long-stay and short-stay patients, we also aimed to capture the length of stay characteristics of intermediate stays patients using correlation analysis. Specifically, a higher correlation between the predicted scores and the actual length of stay indicates better predictive capability. As detailed in Table 2 and the Methods section, our model applied in a zero-shot manner [47] to intermediate-stay patient group showed a significant positive correlation of 0.213 (P-value < 1*e*^-6^) between the predicted probabilities and the actual hospitalization duration. Here the zero-shot manner refers to making predictions for the category of intermediate-stay patients without having previously encountered any data specific to this category. This result was compared with univariate analysis and machine learning-based multivariate analysis. The machine learning methods, trained using five-fold cross-validation on the intermediate-stay patients dataset, exhibited lower correlation, with an average Pearson Correlation Coefficient of 0.179, compared to our model’s average Pearson Correlation Coefficient of 0.213, even in a zero-shot scenario. This suggested that our model effectively captured hospitalization duration characteristics for SCZ patients. Results shown in Figure 4f further supported our finding by categorizing intermediate-stay patients into two groups based on the predicted scores. Patients with predicted scores greater than 0.5 had significantly longer hospitalization durations than those with scores less than 0.5, demonstrating the effectiveness of our model.

**Table 2.**
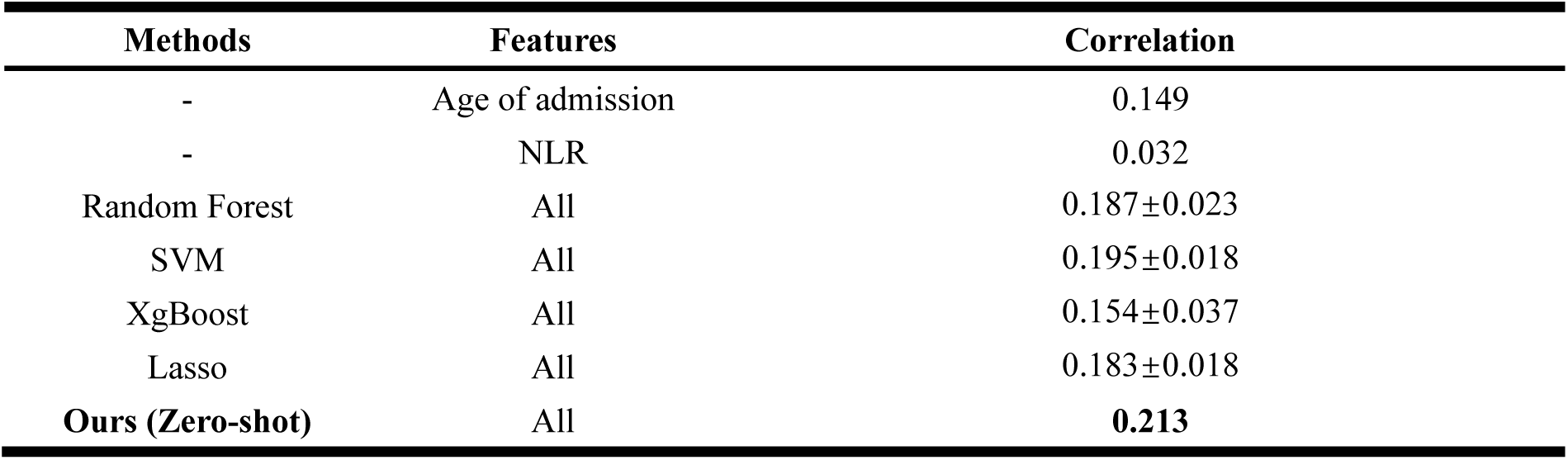
Performance between different models in the population with intermediate length of stay.

| Methods | Features | Correlation |
| --- | --- | --- |
| - | Age of admission | 0.149 |
| - | NLR | 0.032 |
| Random Forest | All | 0.187±0.023 |
| SVM | All | 0.195±0.018 |
| XgBoost | All | 0.154±0.037 |
| Lasso | All | 0.183±0.018 |
| **Ours (Zero-shot)** | All | **0.213** |

## Discussion

This study presented a novel and integrative approach for predicting long-term hospitalization risk in schizophrenia patients by leveraging demographic, behavioral, and blood test indicators using advanced deep learning techniques. Our findings demonstrate the significant potential of using a comprehensive set of admission variables to enhance prediction accuracy and provide deeper insights into the factors influencing prolonged hospitalization in schizophrenia patients.

One of the key contributions of our study is the application of LMs for extracting unstructured information from electronic health records (EHRs). This approach not only ensures high precision and privacy through local deployment but also significantly reduces the manual workload involved in information extraction. The iterative prompt optimization strategy we employed further enhances the model’s accuracy, providing a robust method for extracting relevant behavioral variables with minimal manual intervention. This represents a significant advancement over traditional supervised learning methods. Additionally, we evaluated the effectiveness of our approach against supervised learning methods in two simulated tasks: latent discrimination and erroneous dependency. The results demonstrated that our method significantly reduces the occurrence of both latent discrimination and erroneous dependency issues. This characteristic is particularly crucial in the context of medical-related problems.

Our integrative model achieved a classification accuracy of 0.81 and an AUC of 0.9 on an independent test set, highlighting its efficacy in predicting long-term hospitalization risks. The inclusion of diverse data types, such as demographic variables (e.g., age at admission, disease duration, marital status), blood test indicators, and behavioral data, proved crucial in enhancing the model’s predictive power. This comprehensive approach allows for a more nuanced understanding of the various factors contributing to prolonged hospitalization, supporting better resource allocation and personalized treatment planning at the time of admission.

Our interpretability analysis using SHAP values [41] identified several key risk factors associated with prolonged hospitalization. Age at admission and marital status emerged as significant predictors, consistent with previous findings [18–20]. Behavioral features like self-destructive behaviors and elopement were also important indicators of patient adherence, corroborating earlier research [48]. Additionally, blood test indicators like neutrophil-to-lymphocyte ratio (NLR), lymphocyte percentage, and albumin levels, which have been previously studied in relation to schizophrenia severity and treatment efficacy, were found to be significant predictors in our model [49–51]. This comprehensive analysis not only validates the effectiveness of our model but also provides deeper insights into the factors influencing long-term hospitalization in schizophrenia patients.

Through our model’s interpretability analysis, we identified several risk factors associated with prolonged hospitalization for schizophrenia patients. Some of these risk factors, such as age at admission and marital status, aligned well with established research findings [18–20]. However, other risk factors we identified, such as NLR and lymphocyte, though not previously directly linked to the risk of prolonged hospitalization in schizophrenia, have been associated with the disease severity itself in other studies. For example, behavioral features like self-destructive tendencies and elopement may reflect patient adherence, a relationship corroborated by early research [48]. Blood test indicators, such as NLR, lymphocyte, and albumin have also been previously explored in relation to schizophrenia from perspectives of treatment efficacy and disease severity [49–51]. Given that length of stay is closely tied to the severity of SCZ and the treatment effectiveness, our findings reinforce the relevance of these risk factors. Moreover, our study offers new insights into the relationships between key admission indicators and these critical aspects of schizophrenia management.

Despite the promising results, our study has a few potential limitations. The LM-based information extraction process struggles with long and complex text entries, potentially affecting the accurate capture of relevant behavioral variables. Besides, our dataset, sourced from a single mental health center, may limit the generalizability of our findings. Future studies should validate our model using more diverse, multicenter datasets to enhance its robustness. In addition, due to the limited personal information, our model could not account for confounding factors such as medication adherence, social support, and post-discharge care, which could significantly impact hospitalization outcomes. Future research should include these factors to provide a more comprehensive understanding of the predictors of long-term hospitalization in schizophrenia patients. Despite these limitations, our work provides a valuable new paradigm for mental health record processing and personalized medical guidance. By integrating multi-perspective admission variables, we have significantly enhanced the predictive accuracy and interpretability of long-term hospitalization risks in schizophrenia patients.

In conclusion, our research highlights the effectiveness of using an LM-based paradigm for processing mental health EHRs and incorporating multi-perspective admission variables for long-term hospitalization risk prediction in SCZ patients. This study provides a valuable new paradigm for mental health record processing and personalized medical guidance, contributing to more effective resource allocation and treatment planning at the admission stage.

## Methods

### Data extraction and processing

Our retrospective dataset consists of electronic medical records of SCZ inpatients discharged from the Shanghai Mental Health Center (SMHC) between 2018 and 2022. It includes admission blood test data, demographic information, and current medical history records containing behavioral data. The use of this data was approved by the Ethics Committee of the Shanghai Mental Health Center (Approval No. 2022-79), and strict adherence to data confidentiality regulations was maintained throughout the data processing process.

The data processing and screening process is illustrated in Extended Data Figure 1. Firstly, we excluded patients with a hospitalization duration of less than three days. This is because we considered that a hospitalization duration of less than three days is not typical for acute phase inpatient treatment of SCZ, as these patients may not have received sufficient treatment due to early discharge. Therefore, such a short hospitalization duration does not accurately reflect the treatment efficacy of the patients. Secondly, we excluded patients with outcomes other than cure or improvement upon discharge. These patients may have not yet met the clinical criteria for cure and require extended hospitalization for further treatment, but have been discharged prematurely for non-medical reasons.

The final step involved grouping the records based on hospitalization duration, converting them into binary data representing long-stay and short-stay categories [52]. A study conducted in 2019 indicated that the hospitalization duration for SCZ patients in China is 73 ± 42 days [15]. Building upon this, we defined records with a hospitalization duration exceeding 120 days as long-stay, and those with a duration shorter than 30 days as short-stay. This categorization provided the initial data required for our study.

### Few-shot unstructured information extraction based on local LM

Figure 1a illustrates the workflow of our method. For the selection of local LM, as our dataset is in Chinese, we used Baichuan2-13B [53]. This model has demonstrated performance that is not only close to GPT-3.5 Turbo on various medical question-answering datasets but also comprehensively outperforms LLaMA2 [54] across these test sets. Detailed test result can be found at https://github.com/baichuan-inc/Baichuan2. We downloaded the version released on 2023-12-29 and deployed it on a local server equipped with three Nvidia GeForce 3090 GPUs. For the remote advanced LMs, we used GPT-4 [34], the most advanced model available at the time of our study. Due to external network access restrictions on our local server, we accessed this model via the web interface (accessed on 2024-04-19). We also developed an API-based automated version for users with network access, providing a more automated prompt optimization experience.

Below, we describe our prompt optimization process. First, we annotated 100 examples for each of the three behavioral variables, ensuring a 1:1 ratio of positive to negative samples, meaning there were 50 entries with the corresponding feature and 50 without it. These datasets served as validation sets for adjusting the prompt, which is a hyperparameter for the local LM. Initially, we manually set an initial prompt and had the local LM generate judgments for each entry five times consecutively. If all five judgments were consistent, this was taken as the final judgment; if they were inconsistent, the final judgment was recorded as “not sure.” This process provided an evaluation of the performance of this prompt on the validation set, resulting in counts of correct, incorrect, and uncertain entries. Next, we embedded the current prompt and its corresponding performance into a pre-designed query template and requested the remote LM to optimize the prompt. The optimized prompt then replaced the current prompt. This iterative process continued until no errors were produced on the validation set, and the performance of current prompt ceased to improve after further optimization. Specifically, considering the concentration of information in the text segments we needed to extract, we selected the sentences starting from the two clauses preceding the keyword “impulsion” to the end of the text segment as the model input.

### Comparison of our LM based method with supervised learning-based models

To demonstrate the advantages of our LM-based method over supervised learning-based models under the same conditions, we compared our method with Linear Regression and fine-tuned BERT. Specifically, we annotated 40 entries (20 positive and 20 negative) for each of the three behavioral variables as a test set. The previously annotated validation set of 100 entries was used for training the supervised learning-based models. Linear Regression was implemented using sklearn, and BERT employed the “bert-base-chinese” [55] tokenization strategy and pre-trained parameters. During training, we used a leave-one-out cross validation strategy [39] for hyperparameter tuning.

### Data preprocessing before deep neural network

Before feeding the data into the deep neural network, we followed conventional data preprocessing and encoding steps. For continuous data, we applied a standardization strategy as shown in Equation (1). Specifically, for the duration of illness data, we employed a data binning strategy, dividing the duration into categories of less than 30 days, 30-600 days, and over 600 days, encoded as 0, 0.5, and 1, respectively. Other binary features, such as behavioral variables, were encoded as 0 or 1. Three-category features, such as marital status, were encoded as 0, 0.5, and 1. In order to avoid information leakage, we did standardization after data partitioning, and the standardization parameters of the test set followed the training set.

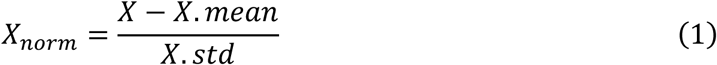

For data partitioning, we employed a time-sliced data splitting approach. This method uses the most recent data in time as an independent test set to simulate the future predictive ability of our model and reduce interference from confounding factors in result evaluation. Here, we designated the last 50 long-term hospitalization records and the last 50 short-term hospitalization records, totaling 100 records, as the independent test set, while the remaining data served as the training set. We did not separately partition a validation set because we utilized leave-one-out cross-validation for hyperparameter selection. This method involves iteratively leaving out one data point as validation data, and by exhaustively enumerating this validation data point, we ultimately obtain the proportion of correct predictions as the evaluation performance on the validation set. The main advantage of this method is its ability to fully utilize the data during training. To expedite computation efficiency, we set the sampling interval of leave-one-out cross-validation to 5, and conducted model training in parallel.

### Prediction model implementation and performance evaluation

As showed in Figure 2a, our model incorporates a self-attention mechanism with the MLP block.

The MLP block can be represented as follows:

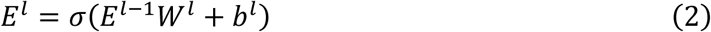

where *E*^l-1^ represents the input vector, *W*^l^ is the trainable weight matrix, *b*^l^ is the bias term, and σ denotes the non-linear activation function. The attention block can be represented as follows:

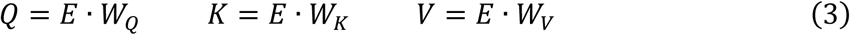

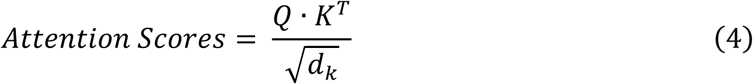

where *E* represents the input vector, *W*_Q_, *W*_K_, and *W*_V_ are trainable weight matrixes. *d*_k_ is the dimension of *K*. Finally, the vectors are weighted summed and fed into a fully connected layer for classification.

We implemented our model using the PyTorch [56] and executed it on a server equipped with three Nvidia GeForce 3090 GPUs. During model training, we employed techniques such as batch normalization [57] and dropout [58]. Cross-entropy was utilized as the loss function, while Adam served as the optimizer [59], with an initial learning rate set to 0.0001 and weight decay to 1e-5. The batch size was set to 32 during training.

We evaluated our model on the independent test set using common machine learning evaluation metrics, as indicated in Supplementary Table 3. To further compare the superiority of our model over machine learning models, we implemented a series of machine learning models using sklearn, also shown in Supplementary Table 3. To ensure fairness, the training process of all machine learning models followed the same procedure as our model.

### Interpretability analysis

We used sklearn to perform Principal Component Analysis dimensionality reduction and visualization analysis on the input vectors and final layer hidden vectors of the test set, using the default parameters. For the SHAP analysis, we used GradientExplainer to calculate the SHAP values. GradientExplainer leverages the gradients of the model output with respect to its input to efficiently approximate SHAP values, particularly suited for deep learning models.

### Ablation study

To validate the effectiveness of multi-perspective admission features, we conducted feature ablation experiments. The data preprocessing methods, dataset partitioning methods, core model architecture, cross-validation methods, and model evaluation schemes used in these experiments were identical to those in the main experiment. The only difference was the change in the dimensionality of the input vectors, which correspondingly altered the input dimension of the first layer of the model architecture.

### Testing on a synthetic external test set

Based on the work of Guevara et al. [60], our synthetic external test set was generated using GPT-4o and reviewed by professional clinicians. The prompts used for generation can be found in the extended data note. To maintain consistency with previous tests, we generated a total of 140 patient history records, with 70 containing negative self-harm information and 70 without. In terms of dataset partitioning, 100 records were allocated for training and validation, while 40 records were reserved for independent testing. The ratio of positive to negative samples in all datasets was kept at 1:1.

### Testing for Bias and Dependency Errors

In our study, we incorporated three types of interference information by generating phrases to be added at the beginning of sentences. All phrases were generated by GPT-4o, with a requirement for diverse categorical expressions. For the dataset addressing racial bias, we created phrases describing Caucasian individuals, such as “A Caucasian patient presents for evaluation,” and phrases for African American individuals, such as “The patient is an African American female.” We conducted two types of tests: the first involved differing phrase types between the training and validation sets, where Caucasian descriptive phrases were included during training and African American phrases during testing. The second test maintained the same phrase types across both sets. This comparison allowed us to analyze the impact of racial bias on the performance of different models. A similar experimental approach was employed for gender bias. For the third type of dependency error testing, we added descriptions related to age, such as “The patient is an elderly female” and “A young patient presents for evaluation,” based on the self-harm negative labels. In this case, our strategy for adding phrases at the beginning of sentences was reversed between the training and testing sets: during training, we added “elderly” phrases to sentences with self-harm indicators and “young” phrases to those without, while the testing set followed the opposite pattern. Clearly, an effective information extraction algorithm should remain unaffected by these irrelevant short phrases.

### Intermediate-stay patients analysis

Intermediate-stay patients are those with hospitalization durations ranging from 30 to 120 days. After applying the same screening criteria as in the main experiment, a total of 3,916 records were included. We chose to use the Spearman correlation coefficient to measure performance because it focuses more on the direct monotonic relationship between variables. For the machine learning models, we used their regression algorithm versions and trained them on the intermediate-stay patients dataset using five-fold cross-validation for the regression task. All feature extraction and data preprocessing methods were consistent with those used in the main experiment.

## Code availability

Source code can be found at https://github.com/RoarBoil/SchizoLOSPrediction

## Data Availability

All data produced in this study can be obtained from the authors upon reasonable request, with the approval of the Shanghai Mental Health Center Ethics Committee.

Due to ethical restrictions, the data used in this study cannot be publicly shared. For further inquiries about the data, please contact the corresponding author. Data access can be granted under the following conditions: (1) all co-authors agree to share the data; (2) there is a data use agreement and ethical approval in place, following the data application requirements of the Shanghai Mental Health Center. Once shared, the data can only be used for the specified purpose.

## Acknowledgments

This work was supported by grants from STI 2030—Major Projects (no. 2022ZD0209100), the National Natural Science Foundation of China (nos. 81971292 and 82150610506), the Natural Science Foundation of Shanghai (no. 21ZR1428600), the Medical-Engineering Cross Foundation of Shanghai Jiao Tong University (nos. YG2022ZD026 and YG2023ZD27), and SJTU Trans-med Awards Research (no. 20220103)

## Ethics statement

This retrospective study was approved by the Ethics Committee of Shanghai Mental Health Center (approval number 2022-79).

## Conflict of interest

The authors declare no competing interests.

## Extended Data Figure

**Extended Data Figure 1.**
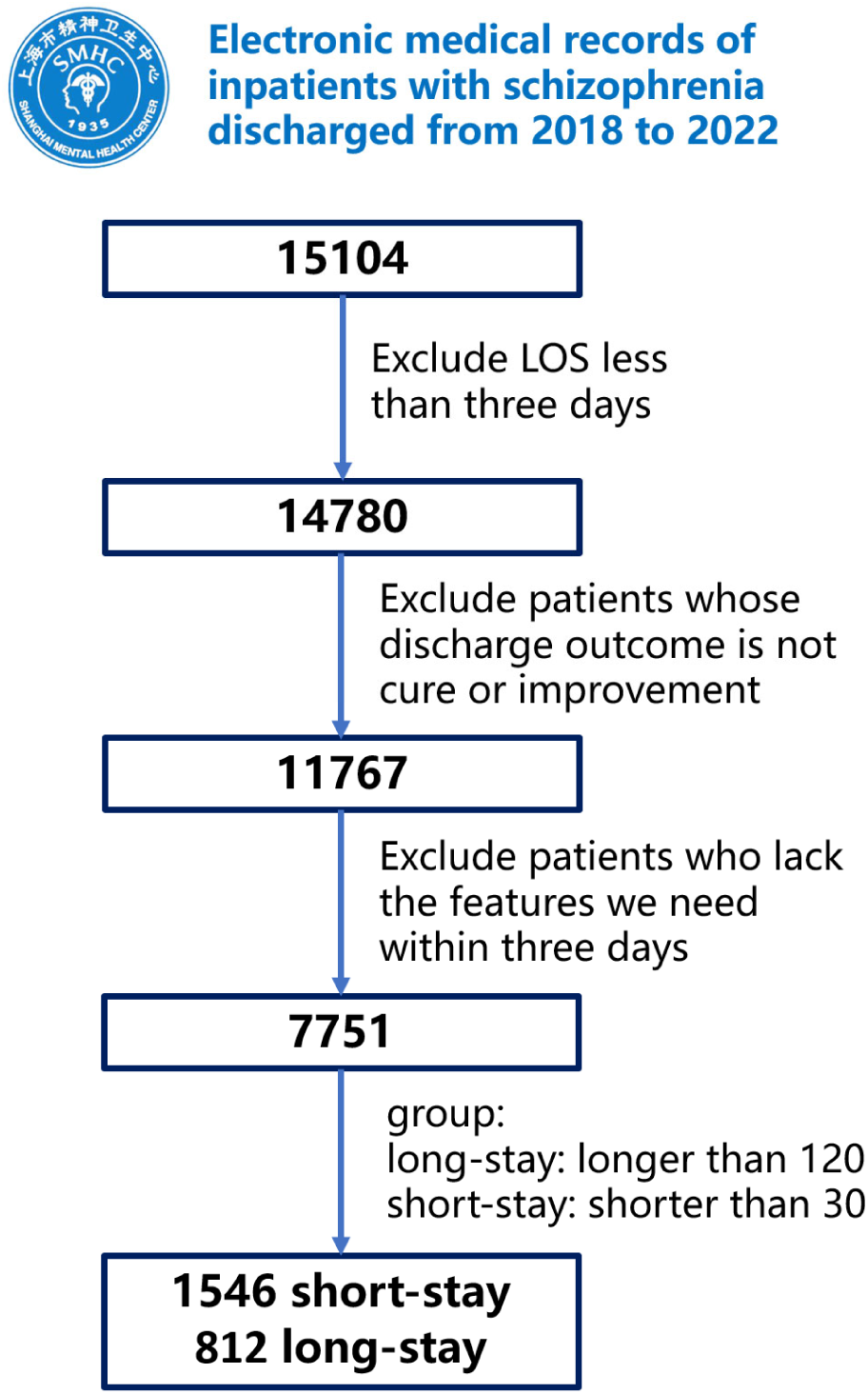
Data screening and preprocessing process in our study.

**Extended Data Figure 2.**
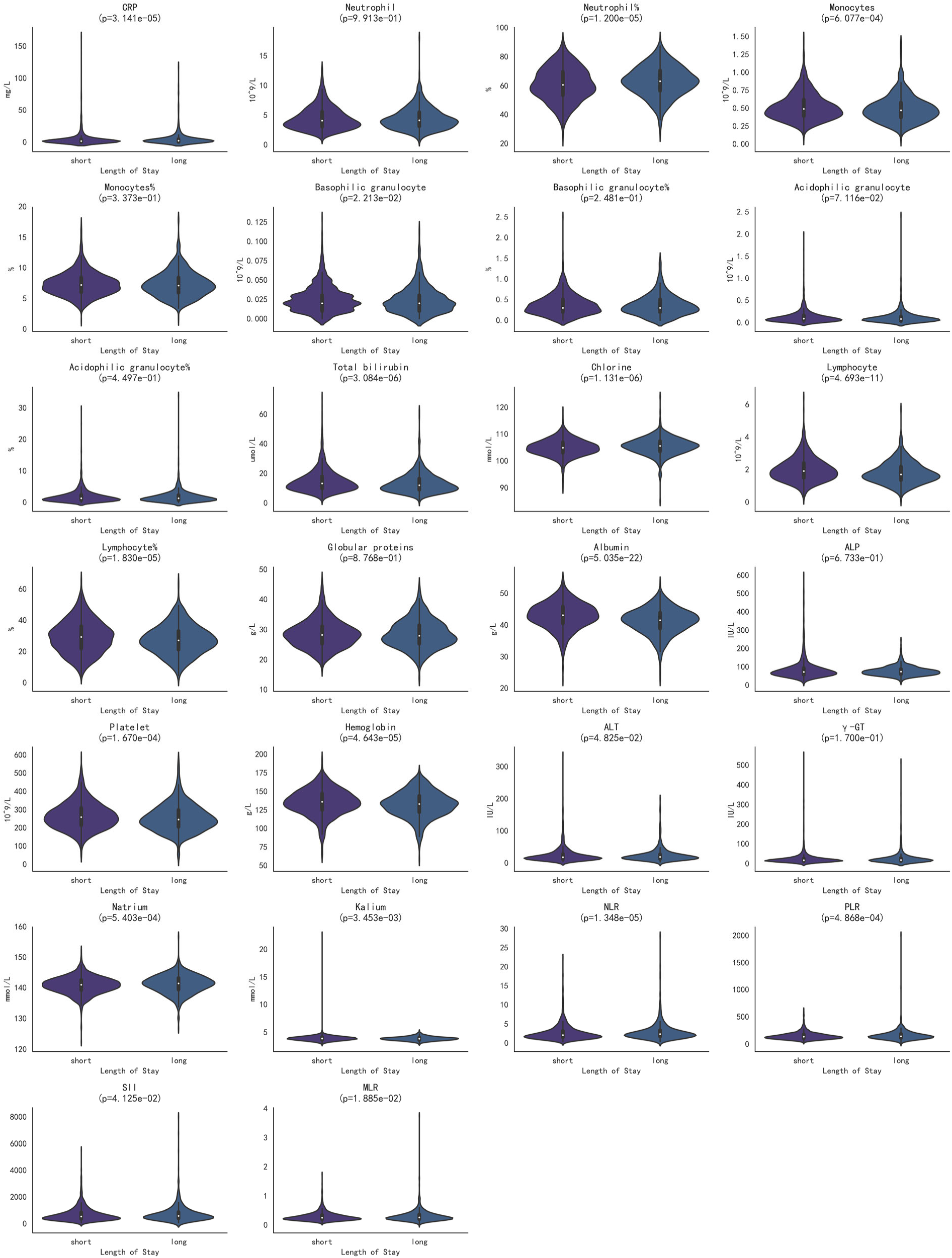
Violin plot of blood test variables. Significance testing using the Mann-Whitney U test

**Extended Data Figure 3.**
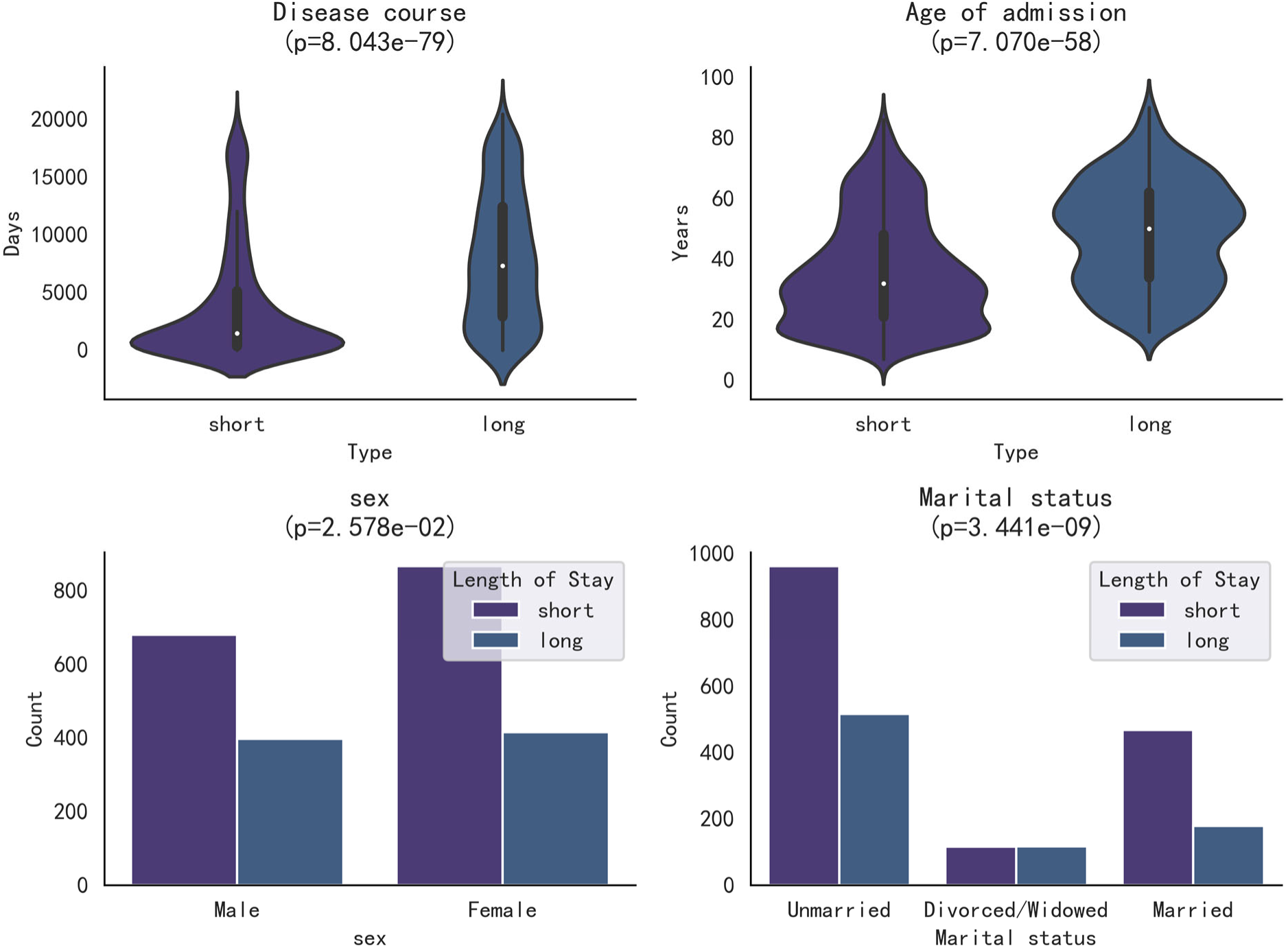
Violin plot of demographic variables. Significance testing using the Mann-Whitney U test for continuous variables and chi-square test of homogeneity for categorical variables.

**Extended Data Figure 4.**
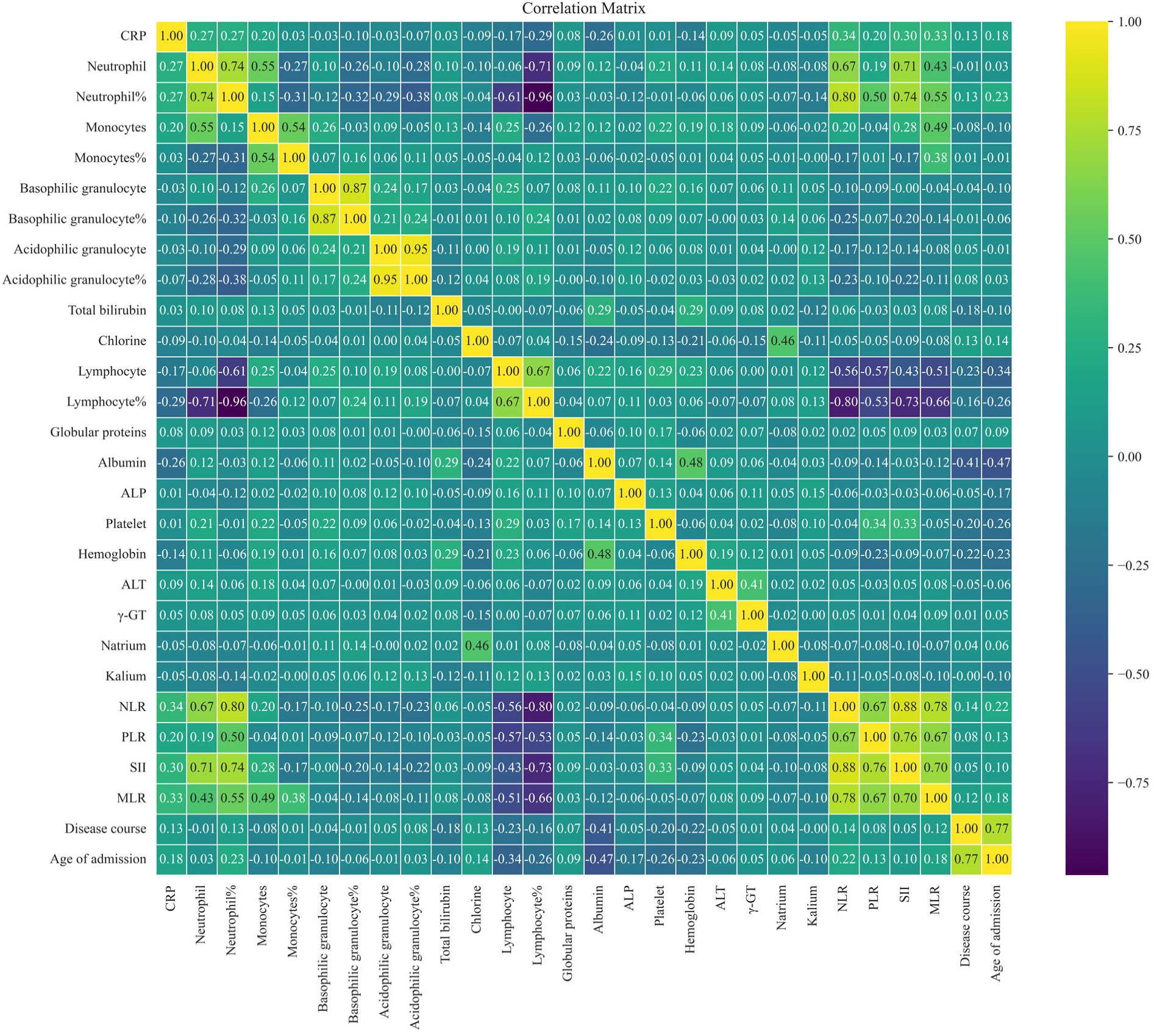
Correlation analysis between continuous variables.

**Extended Data Figure 5.**
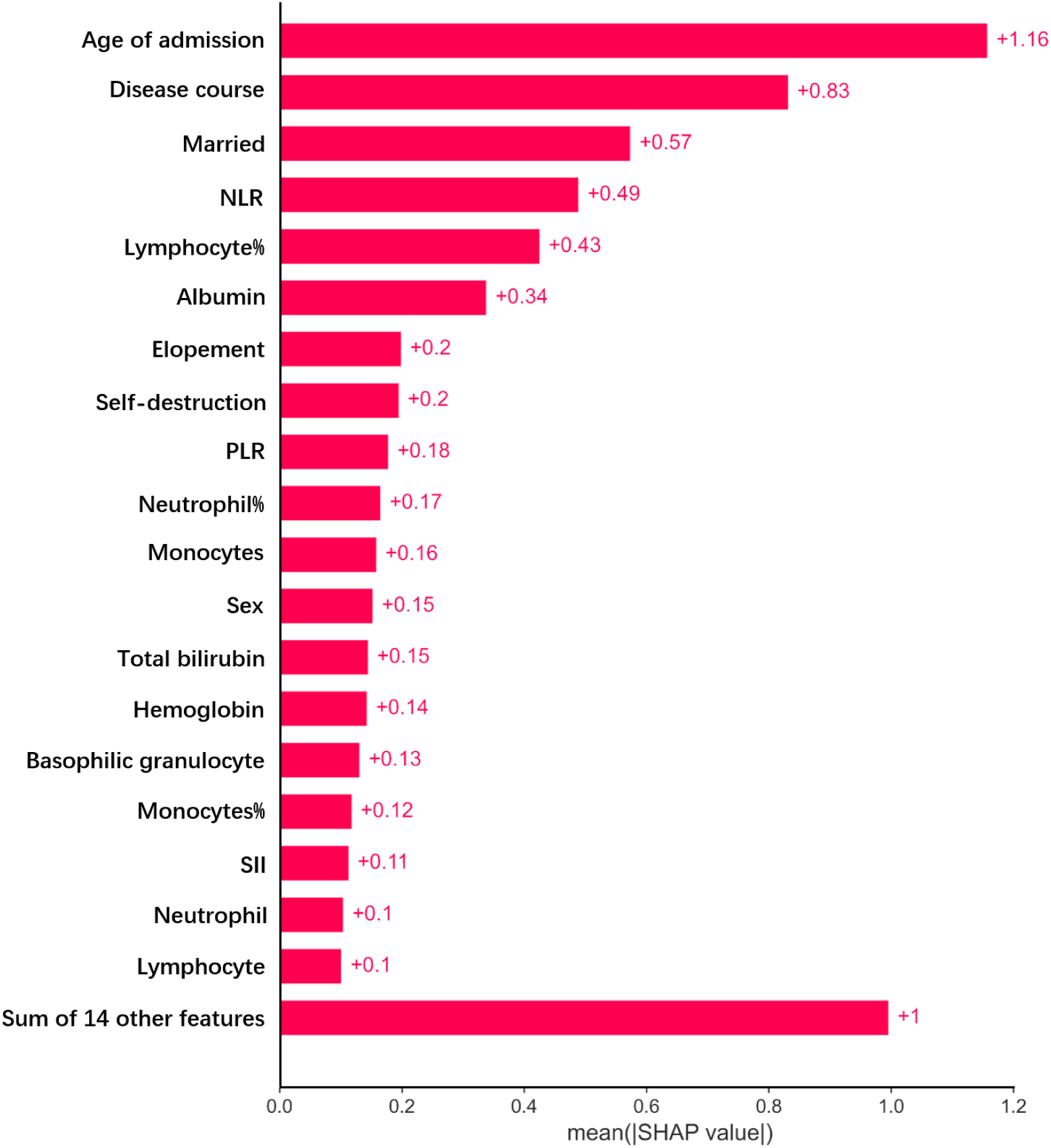
Feature importance analysis using SHAP

**Extended Data Figure 6.**
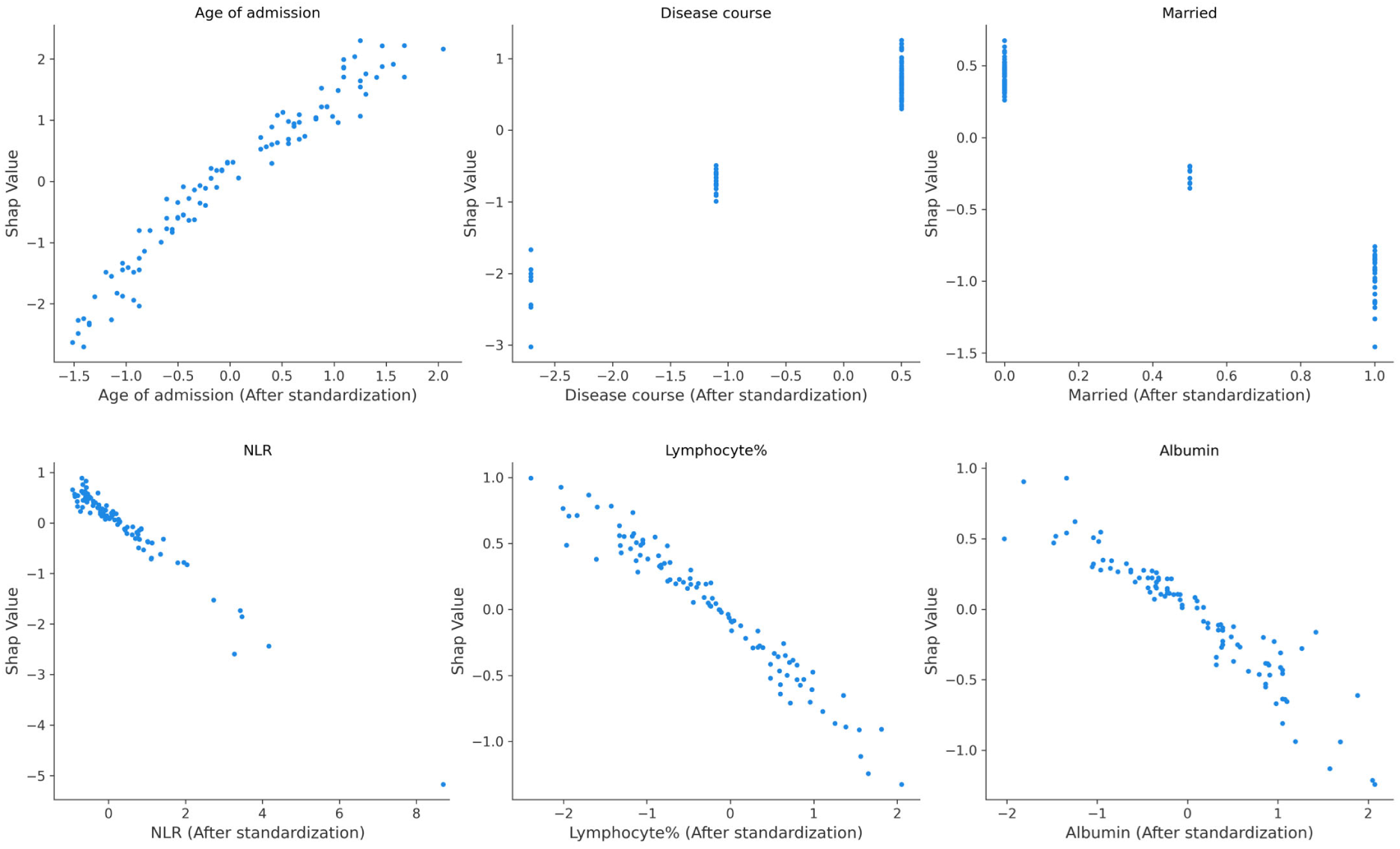
Dependency plot of the three most important demographic variables and the three most important blood test indicators.

**Extended Data Figure 7.**
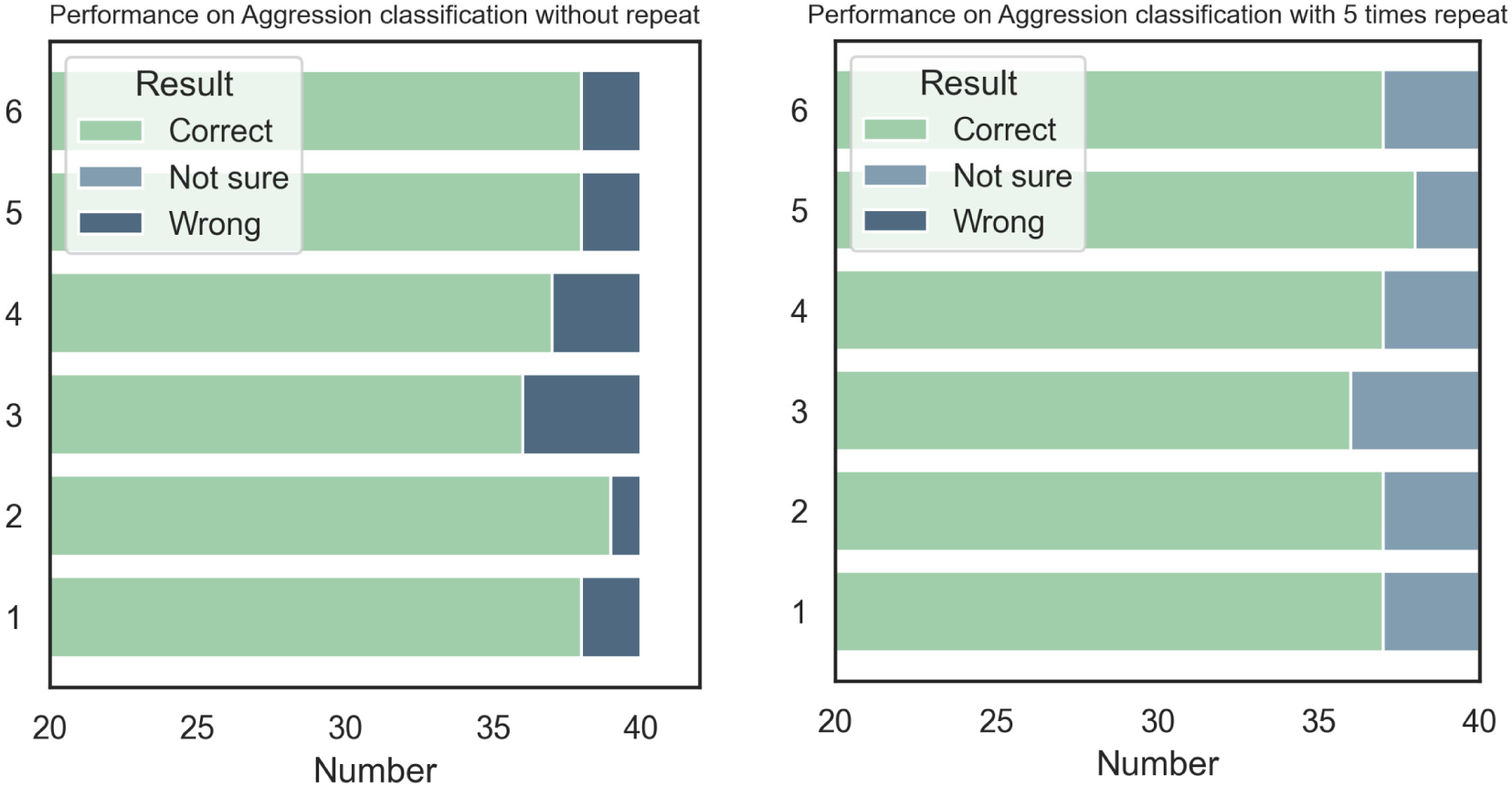
The analysis of the LM paradigm across varying repetition counts indicates that the five-repetition paradigm exhibits significantly higher stability and a lower error rate than the paradigm without repetition.

**Extended Data Figure 8.**
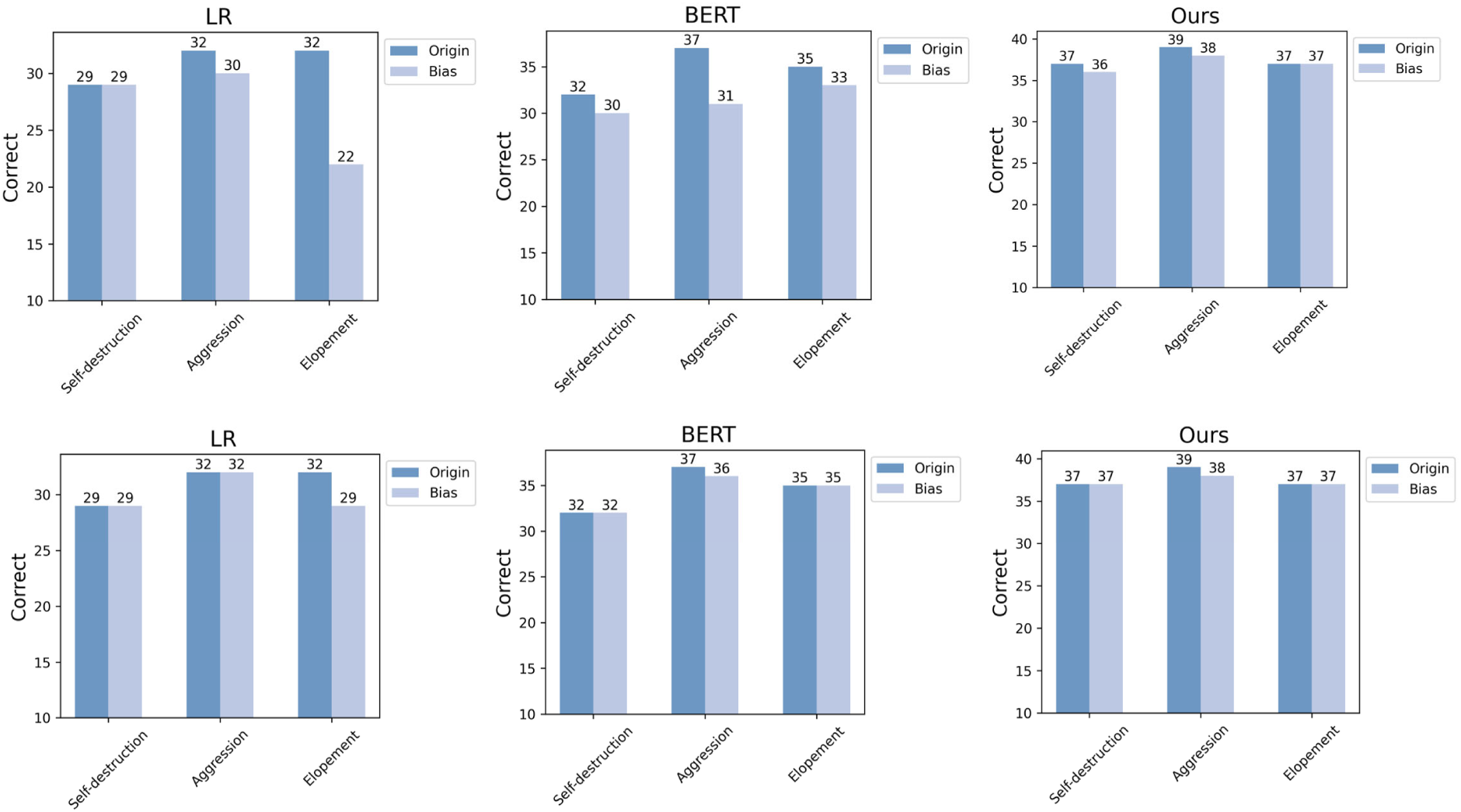
The test results on racial bias are presented, with the first row indicating the outcomes under conditions where bias exists in both the training and testing datasets, while the second row reflects the results when there is no bias in either dataset.

**Extended Data Figure 9.**
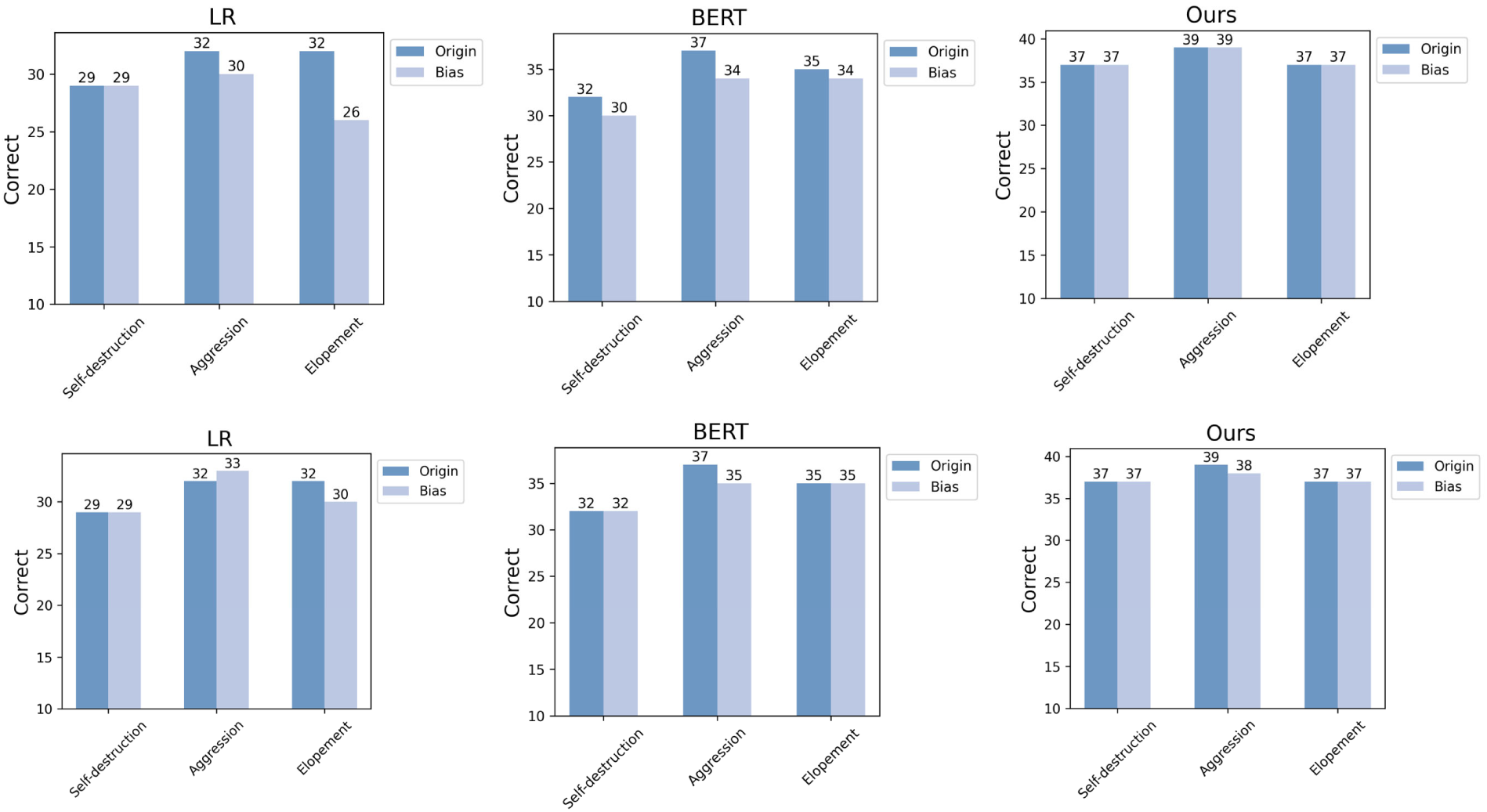
The test results on gender bias are presented, with the first row indicating the outcomes under conditions where bias exists in both the training and testing datasets, while the second row reflects the results when there is no bias in either dataset.

**Extended Data Figure 10.**
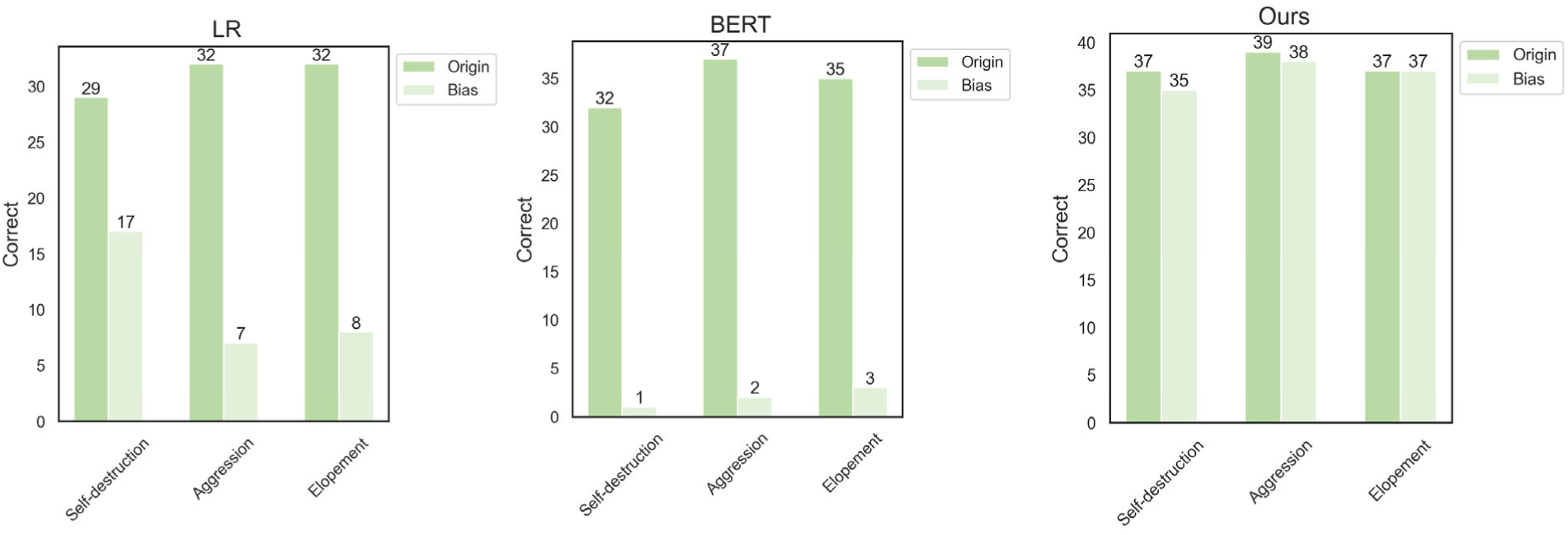
The test results on dependency error are presented.

## Extended Data Note

### Definition of behavioral variables

In this study, we defined three important behavioral variables that are considered to be highly related to the length of hospital stay.

1. Aggression: Any intention or action characterized by destructiveness and impulsiveness.
2. Self-destruction: Any ideation or behaviors involving suicide or self-injury.
3. Elopement: Any attempt to escape from home, hospital, or other healthcare facility.

### Examples of unstructured information

After using the preprocessing methods described in Methods, the following are examples of input data that need to be extracted using the extraction method we developed.

1. Urination and defecation are normal. Denies negative behaviors but has elopement or impulsive actions.
2. Recently has had constipation, sleep is generally okay, but easily startled. Has impulsive behaviors such as waving knives, but no negative or elopement behavior.
3. Urinates on themselves when motionless and silent, with no significant weight changes. No impulse to harm others, suicidal tendencies, self-harm, or elopement.
4. Urination and defecation are normal, with no significant weight changes. Exhibits clear impulsive behaviors of property destruction, denies elopement or negative behaviors.

### Prompt Example for Generating Synthetic Datasets

Please assist me in generating 10 entries for the admission history of electronic medical records, specifically focusing on the current medical history of patients with schizophrenia. Each entry should include a description of whether the patient has a history of self-injurious behavior (without directly using the term “self-injury”). The types of entries should vary as much as possible, ensuring that 5 entries indicate a history of such behavior and 5 entries do not.

### Detailed blood test indicators analysis in case study

The predictive test indicator characteristics are primarily divided into three categories: blood cell and ratio-related indicators, liver function-related indicators, and electrolyte-related indicators.

From the perspective of blood cell-related indicators, the patient exhibits a slightly lower than normal lymphocyte ratio, a higher-than-normal neutrophil and basophil ratio, and an elevated monocyte ratio, with varying predictive directions for these features. Previous studies have found that schizophrenia is associated with elevated white blood cell counts, with high inflammation characteristics linked to increased permeability of the blood-brain barrier to inflammatory cells, also suggesting the possibility of severe cognitive deficits. This does not fully align with our model’s explanation, but it is important to consider that changes in blood cell indicators represent a complex process, and a single indicator may be insufficient to fully explain the symptoms of the disease. The predictive contradictions among blood cell test indicators may suggest that we need to view cell changes as an overall indicator rather than isolating each indicator. During the acute phase in schizophrenia patients, increased monocytes in cerebrospinal fluid and activation of monocyte macrophages are associated with paranoid schizophrenia. A higher monocyte ratio may indicate the severity of the disease. Our model includes peripheral blood indicators that are easy to detect, although these indicators do not directly suggest central nervous system blood markers. However, existing research has confirmed a correlation between central and peripheral blood markers, although the specific connection remains unclear, indicating the potential of peripheral blood markers in predicting clinical symptoms. This also aligns with the finding that a high monocyte ratio in this patient may serve as a predictive factor for long-term hospitalization. Additionally, studies have suggested that immune-inflammatory indicators in persistent paranoid schizophrenia patients are lower than in episodic paranoid schizophrenia patients. Persistent symptoms are typically associated with treatment difficulties and longer hospitalization periods. This may explain why a lower lymphocyte ratio plays an important role in predicting long-term hospitalization in this case. The differences in predictive directions for other cell indicators may also be due to this reason. Although research conclusions on full blood cell count inflammatory markers are often contradictory, they generally support the view that chronic inflammation is a significant biological characteristic of schizophrenia. In our study, elevated PLR is an important marker for long-term hospitalization, possibly representing the progression of the acute phase.

Regarding liver function-related indicators, the patient’s alkaline phosphatase and albumin levels are slightly higher than normal, while total bilirubin and globulin levels are slightly lower than normal. Bilirubin has been proven to have anti-inflammatory effects, and lower total bilirubin levels may be associated with cognitive impairment in the patient. Liver function impairment plays a significant role in the classification of psychiatric disorders. Elevated alkaline phosphatase may be related to metabolic abnormalities in schizophrenia patients, leading to increased blood glucose and lipid levels, thereby increasing cardiovascular disease risk. In this case, the patient’s cranial MRI shows multiple ischemic foci in the frontal and parietal lobes, indicating cardiovascular disease risk, which is one of the key factors associated with increased early mortality and reduced life expectancy in schizophrenia patients and is significantly associated with long-term hospitalization. Metabolic phenotype abnormalities may indicate this cardiovascular risk. Additionally, decreased globulin and albumin levels are associated with depression risk in schizophrenia patients and are also risk factors for schizophrenia. Studies have confirmed a negative correlation between hemoglobin concentration and PANSS scores, with tissue hypoxia leading to functional disorders and poor prognosis. Although the patient’s hemoglobin level is slightly higher than normal, which theoretically should be a protective factor, other risk factors overshadow this protective effect. Therefore, while higher hemoglobin tends to predict shorter hospitalization duration, this predictive effect is weakened due to the presence of other risk factors.

In terms of electrolyte-related indicators, the patient’s chloride ion concentration is slightly lower than normal. Chloride ion concentration determines the activation and polarity of GABA, with excitatory GABA promoting neuron differentiation; premature or delayed maturation may lead to abnormalities. Reviewing the patient’s medication orders reveals that the patient was only administered olanzapine during this hospitalization. As a muscarinic receptor antagonist, olanzapine may affect calcium ion channels and, subsequently, calcium-dependent chloride ion concentration. Olanzapine may also induce metabolic disorders through multiple signaling pathways, leading to poor prognosis and increased risk of long-term hospitalization, which aligns with the model’s prediction.

---

**Supplementary Table 1.** Changes in model performance during prompt optimization process.

| Task | Result | Round1 | Round2 | Round3 | Round4 | Round5 |
| --- | --- | --- | --- | --- | --- | --- |
| Self-destruction | wrong | 9 | 2 | 0 | 0 | - |
|  | not sure | 32 | 5 | 6 | 5 | - |
|  | correct | 59 | 93 | 94 | 95 | - |
| Aggression | wrong | 1 | 0 | 2 | 0 | 0 |
|  | not sure | 16 | 57 | 1 | 5 | 4 |
|  | correct | 83 | 43 | 97 | 95 | 96 |
| Elopement | wrong | 3 | 1 | 2 | 0 | - |
|  | not sure | 22 | 6 | 1 | 5 | - |
|  | correct | 75 | 93 | 97 | 95 | - |
| Synthetic | wrong | 2 | 0 | 0 | - | - |
|  | not sure | 11 | 5 | 2 | - | - |
|  | correct | 87 | 95 | 98 | - | - |

**Supplementary Table 2.** Performance comparison on test set.

| Task | Result | Linear Regression | BERT-finetuned | Ours |
| --- | --- | --- | --- | --- |
| Self-destruction | wrong | 11 | 8 | 0 |
|  | not sure | 0 | 0 | 3 |
|  | correct | 29 | 32 | 37 |
| Aggression | wrong | 8 | 3 | 0 |
|  | not sure | 0 | 0 | 1 |
|  | correct | 32 | 37 | 39 |
| Elopement | wrong | 8 | 5 | 0 |
|  | not sure | 0 | 0 | 3 |
|  | correct | 32 | 35 | 37 |
| Synthetic | wrong | 8 | 3 | 0 |
|  | not sure | 0 | 0 | 2 |
|  | correct | 32 | 37 | 38 |

**Supplementary Table 3.**
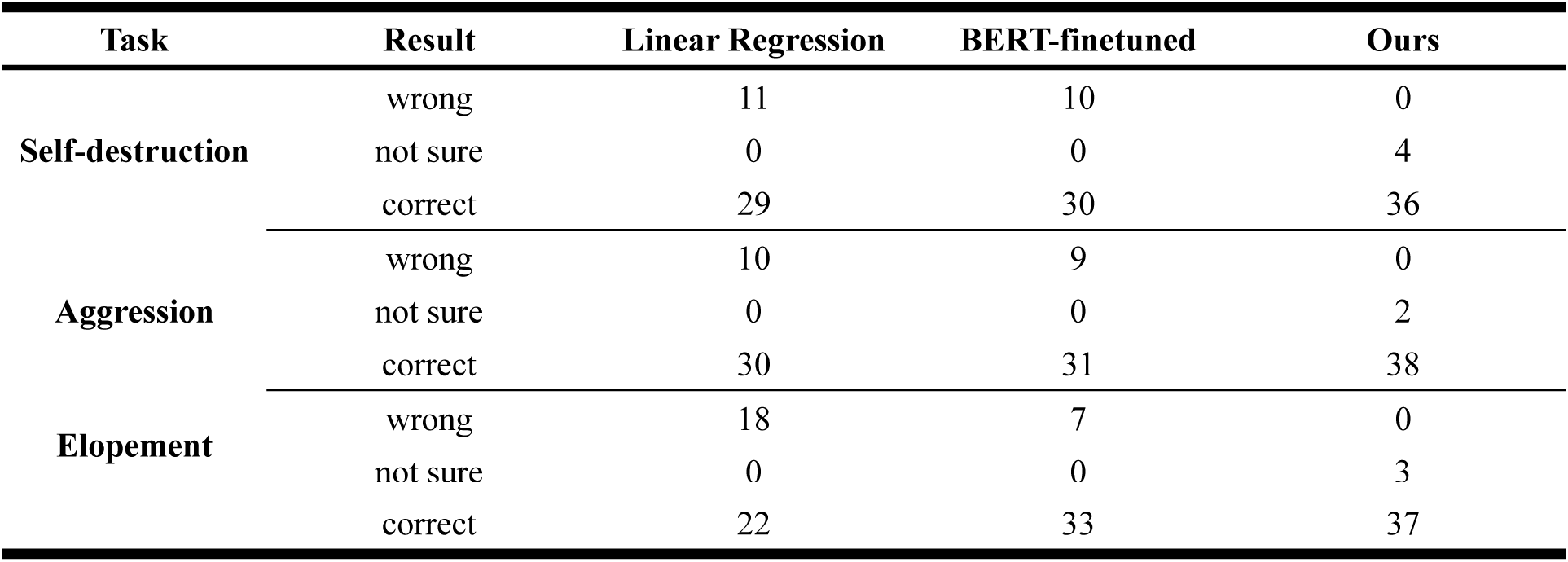
Bias test result (White/Black)

**Supplementary Table 3.** Bias test result (White/White)

| Task | Result | Linear Regression | BERT-finetuned | Ours |
| --- | --- | --- | --- | --- |
| Self-destruction | wrong | 11 | 8 | 0 |
|  | not sure | 0 | 0 | 3 |
|  | correct | 29 | 32 | 37 |
| Aggression | wrong | 8 | 4 | 0 |
|  | not sure | 0 | 0 | 2 |
|  | correct | 32 | 36 | 38 |
| Elopement | wrong | 11 | 5 | 0 |
|  | not sure | 0 | 0 | 3 |
|  | correct | 29 | 35 | 37 |

**Supplementary Table 4.** Bias test result (Male/Female)

| Task | Result | Linear Regression | BERT-finetuned | Ours |
| --- | --- | --- | --- | --- |
| Self-destruction | wrong | 11 | 10 | 0 |
|  | not sure | 0 | 0 | 3 |
|  | correct | 29 | 30 | 37 |
| Aggression | wrong | 10 | 6 | 0 |
|  | not sure | 0 | 0 | 1 |
|  | correct | 30 | 34 | 39 |
| Elopement | wrong | 14 | 6 | 0 |
|  | not sure | 0 | 0 | 3 |
|  | correct | 26 | 34 | 37 |

**Supplementary Table 5.** Bias test result (Male/ Male)

| Task | Result | Linear Regression | BERT-finetuned | Ours |
| --- | --- | --- | --- | --- |
| Self-destruction | wrong | 11 | 8 | 0 |
|  | not sure | 0 | 0 | 3 |
|  | correct | 29 | 32 | 37 |
| Aggression | wrong | 7 | 5 | 0 |
|  | not sure | 0 | 0 | 2 |
|  | correct | 33 | 35 | 38 |
| Elopement | wrong | 11 | 5 | 0 |
|  | not sure | 0 | 0 | 3 |
|  | correct | 30 | 35 | 37 |

**Supplementary Table 6.** Dependency error test.

| Task | Result | Linear Regression | BERT-finetuned | Ours |
| --- | --- | --- | --- | --- |
| Self-destruction | wrong | 23 | 39 | 0 |
|  | not sure | 0 | 0 | 5 |
|  | correct | 17 | 1 | 35 |
| Aggression | wrong | 33 | 38 | 0 |
|  | not sure | 0 | 0 | 2 |
|  | correct | 7 | 2 | 38 |
| Elopement | wrong | 32 | 37 | 0 |
|  | not sure | 0 | 0 | 3 |
|  | correct | 8 | 3 | 37 |

**Supplementary Table 7.** Performance evaluation of multiple models on independent test sets.

| Model | Accuracy | Precision | Recall | F1 | MCC |
| --- | --- | --- | --- | --- | --- |
| Random Forest | 0.69 | 0.81 | 0.5 | 0.62 | 0.41 |
| XGBoost | 0.68 | 0.71 | 0.60 | 0.65 | 0.36 |
| SVM | 0.69 | 0.77 | 0.54 | 0.64 | 0.40 |
| Lasso | 0.68 | 0.78 | 0.50 | 0.61 | 0.39 |
| **Ours** | **0.81** | **0.83** | **0.78** | **0.80** | **0.62** |

**Supplementary Table 8.** Performance evaluation for ablation study.

| Model | Accuracy | Precision | Recall | F1 | MCC |
| --- | --- | --- | --- | --- | --- |
| Demographic | 0.73 | 0.71 | 0.78 | 0.74 | 0.46 |
| Behavioral + Demographic | 0.74 | 0.79 | 0.66 | 0.72 | 0.49 |
| Blood + Demographic | 0.77 | 0.78 | 0.76 | 0.77 | 0.54 |
| **Full** | **0.81** | **0.83** | **0.78** | **0.80** | **0.62** |

## Notes

### Competing Interest Statement

The authors have declared no competing interest.

### Author Declarations

Ethics committee of Shanghai Mental Health Center gave ethical approval for this work (Approval Letter No.2022-79)

